# Meta-analysis of the amyotrophic lateral sclerosis spectrum uncovers genome instability

**DOI:** 10.1101/2022.08.11.22278516

**Authors:** Oliver J. Ziff, Jacob Neeves, Jamie Mitchell, Giulia Tyzack, NeuroLINCS consortium, AnswerALS consortium, Carlos Martinez Ruiz, Nicholas McGranahan, Raphaelle Luisier, Anob M. Chakrabarti, Simon J. Boulton, Gavin Kelly, Jack Humphrey, Rickie Patani

**Affiliations:** The Francis Crick Institute, 1 Midland Road, London NW1 1AT, UK; Department of Neuromuscular Diseases, Queen Square Institute of Neurology, University College London, WC1N 3BG, UK; National Hospital for Neurology and Neurosurgery, University College London NHS Foundation Trust, WC1N 3BG, UK; Cancer Genome Evolution Research Group, Cancer Research UK Lung Cancer Centre of Excellence, University College London Cancer Institute, London, UK; Genomics and Health Informatics Group, Idiap Research Institute, Martigny, Switzerland; Nash Family Department of Neuroscience & Friedman Brain Institute, Icahn School of Medicine at Mount Sinai, New York, NY, USA

## Abstract

Amyotrophic Lateral Sclerosis (ALS) is characterised by progressive motor neuron degeneration but there is marked genetic and clinical heterogeneity^1^. Identifying common mechanisms of ALS amongst this diversity has been challenging, however, a systematic framework examining motor neurons across the ALS spectrum may reveal unifying insights. Here, we present the most comprehensive compendium of ALS human-induced pluripotent stem cell-derived motor neurons (iPSNs) from 429 donors across 15 datasets including Answer ALS and NeuroLINCS, spanning 10 ALS mutations and sporadic ALS. Using gold-standard reproducible bioinformatic workflows, we identify that ALS iPSNs show common activation of the DNA damage response and p53 signalling, which was replicated in the NYGC ALS postmortem cohort of 203 spinal cord samples. The strongest p53 activation was observed in *C9orf72* repeat expansions but was also independently increased in *TARDBP, FUS* and sporadic subgroups. ALS iPSNs showed extensive splicing alterations and enrichment of SNVs, indels and gene fusions, which may contribute to their damage-induced mutation signature. Our results integrate the global landscape of motor neuron alterations in ALS, revealing that genome instability is a common hallmark of ALS motor neurons and provides a resource to identify future ALS drug targets.

## Introduction

Over 30 gene mutations have been established to cause ALS, the most common being in *C9orf72, SOD1, TARDBP* and *FUS*. However, in ∼90% of cases, no pathogenic mutation is identified, which we refer to as sporadic (sALS)^1^. Heritability estimates of ALS are ∼50% and genome-wide association studies have identified over 50 common variants associated with ALS susceptibility (e.g. *UNC13A, TBK1, ATXN2, NEK1, SMN1*)^2^. These genes are involved in a wide range of processes spanning neuronal functions, RNA processing, DNA damage response, mitochondrial function and proteostasis^3^. This genomic complexity has made identifying common pathogenic mechanisms across ALS challenging^4^.

A major hurdle in identifying the causes of ALS has been the inaccessibility to patient motor neurons. Whilst postmortem ALS tissue has revealed important insights, it represents the end-stage of the disease with only few surviving motor neurons^5–8^. Human-induced pluripotent stem cell (iPSC)-derived motor neurons (iPSNs) offer a potential solution. They can recapitulate pathological features of ALS, enabling exploration of the functional consequences of genetic variants on motor neurons during the initial phases of the disease^9–11^. Since iPSNs can be generated from any individual irrespective of genetic background, they enable sALS to be modelled, which is not possible with animal models^12^. However, iPSN cultures are expensive and labour intensive and studies have often been limited to 3 or fewer patients^13–17^. Against this background, there has been a recent expansion of ALS iPSN biobanks with initiatives such as neuroLINCS^18^ and Answer ALS^19^, offering a unique opportunity to identify generalisable motor neuron perturbations across ALS genetic backgrounds. Here, we present a robust analytical framework to identify unifying molecular aberrations underlying motor neuron dysfunction in ALS.

## Results

### iPSC-derived motor neuron identities

Our search strategy of databases identified 16 ALS iPSN datasets that had undergone RNA-sequencing (RNA-seq; Fig. 1, Extended Data Fig. 1). iPSN differentiation protocols for each dataset were extracted and found to follow generally similar procedures between datasets, however, there were notable differences in the duration of cultures, which ranged between 12-42 days *in vitro* (mean 31 days; Table S1). All samples underwent extensive quality control and principal component analysis was used to interrogate the effects of sequencing and culture batch confounding variables (Table S2). This revealed 2 global clusters of iPSNs separated by PC1 and PC2, according to poly(A) or total Ribo-Zero RNA library preparation (Extended Data Fig. 2-3). The principal component gene loadings confirmed that the separation was driven by histone and small nucleolar encoding genes, which represent non-polyadenylated genes (Extended Data Fig. 2a).

**Fig. 1:**
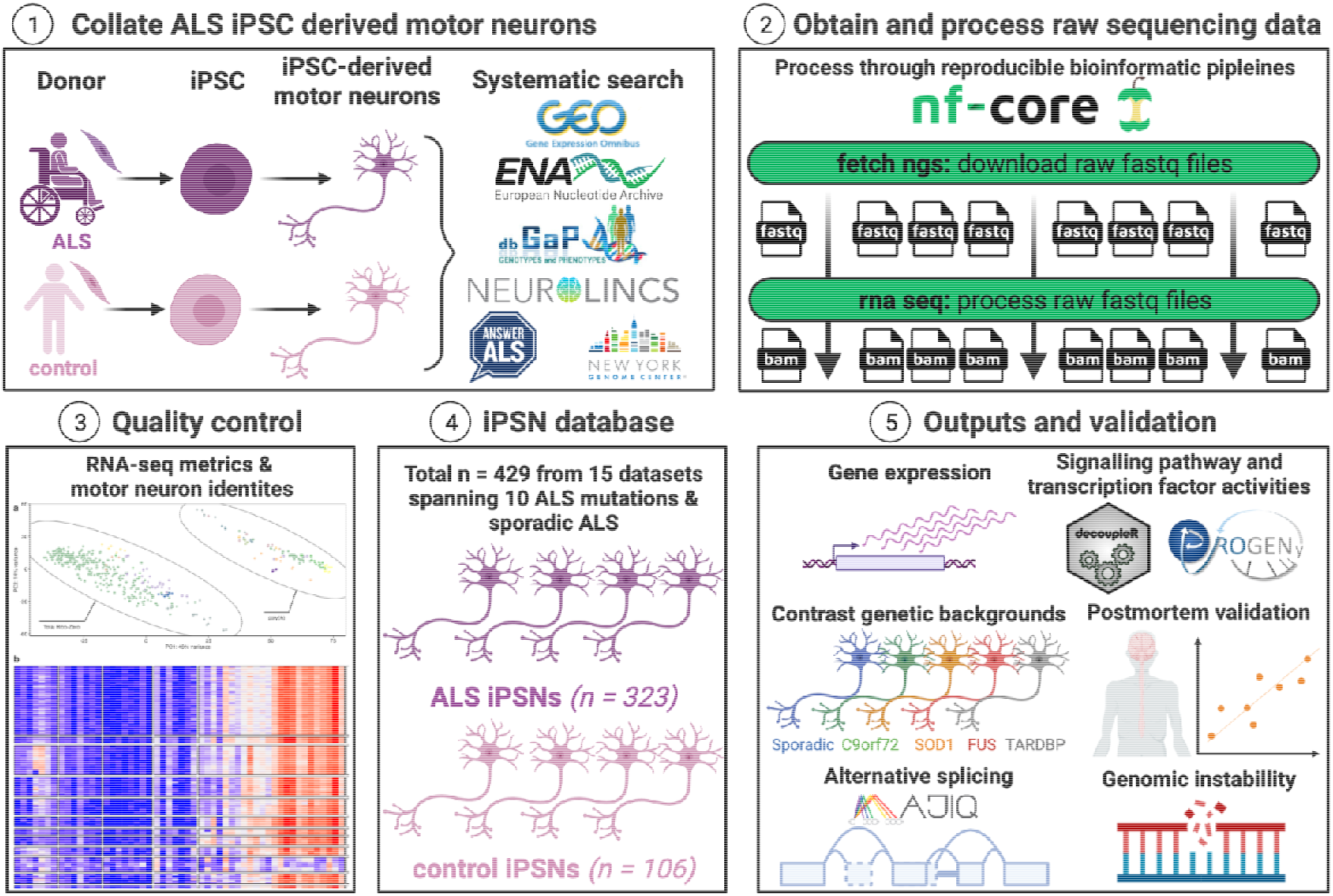
Study overview. Schematic summarising our analytic framework using iPSC-derived motor neurons (iPSNs) to interrogate perturbations across the spectrum of ALS.

iPSNs showed high expression of neuronal markers from all datasets with exception of one HB9 reporter dataset that also exhibited different RNA library preparations between ALS (Ribo-Zero) and control (polyA) samples and was excluded from the meta-analysis (Extended Data Fig. 4)^20^. Although the expression of post-mitotic dorso-ventral motor neuron domain markers (e.g. *CHAT, MNX1 [HB9], LHX3, FOXP1, ALDH1A2, ISL1*) varied between datasets, there was high expression of rostro-caudal markers (*HOX1-8)* across all datasets, which is consistent with hindbrain, cervical and thoracic spinal cord specification (Extended Data Fig. 5-7). For the meta-analysis, we included 15 datasets comprising 429 iPSNs, of which 323 were from ALS patients and 106 from non-ALS controls. ALS iPSNs carried pathogenic mutations in 10 different genes, including *C9orf72*^*18,19,21–24*^ (n = 60), *SOD1*^*14,15,17–19*^ (n = 20), *FUS*^*16,19,21,25,26*^ (n = 14) and *TARDBP*^*19,22,27,28*^ (n = 10), whilst 208 (64.2%) were from patients without an identifiable ALS mutation, which we term sALS (Table 1).

**Table 1.** iPSN datasets included in meta-analysis.

| Reference | Accession # | Mutation | ALS n | Control n | Library type | Layout |
| --- | --- | --- | --- | --- | --- | --- |
| Sareen et al, 2013 | GSE52202 | C9orf72 | 4 | 4 | polyA | Single |
| Kiskinis et al, 2014 | GSE54409 | SOD1 | 2 | 3 | polyA | Paired |
| Kapeli et al, 2016 | GSE77702 | FUS | 3 | 2 | polyA | Single |
| Wang et al, 2017 | GSE95089 | SOD1 | 2 | 2 | polyA | Paired |
| De Santis et al, 2017 | GSE94888 | FUS | 3 | 3 | Ribo-zero | Paired |
| Bhinge et al, 2017 | PRJNA361408 | SOD1 | 2 | 2 | Ribo-zero | Single |
| Luisier et al, 2018 | GSE98290 | VCP | 3 | 3 | polyA | Single |
| Abo-Rady et al, 2020 | GSE143743 | C9orf72 | 3 | 3 | polyA | Single |
| Dafinca et al. 2020 | GSE139144 | C9orf72 | 4 | 8 | polyA | Paired |
|  | GSE147544 | TARDBP | 6 | 4 |  |  |
| Catanese et al., 2021 | GSE168831 | C9orf72 | 6 | 6 | polyA | Paired |
|  |  | FUS | 6 |  |  |  |
| Smith et al, 2021 | PRJEB47567 | TARDBP | 3 | 2 | polyA | Paired |
| Hawkins et al, 2022 | GSE203168 | FUS | 2 | 2 | Ribo-zero | Single |
| Sommer et al, 2022 | GSE201407 | C9orf72 | 6 | 6 | polyA | Paired |
| NeuroLINCS, 2022 | phs001231.v2.p1 | sporadic | 8 | 14 | Ribo-zero | Paired |
|  |  | SOD1 | 6 |  |  |  |
|  |  | C9orf72 | 16 |  |  |  |
| Answer ALS, 2022 | AnswerALS data portal | sporadic | 200 | 42 | Ribo-zero | Paired |
|  |  | C9orf72 | 21 |  |  |  |
|  |  | SOD1 | 8 |  |  |  |
|  |  | 6 other ALS mutations | 9 |  |  |  |
| Total |  |  | 323 | 106 |  |  |

### ALS iPSNs activate the DNA damage response

To identify pan-ALS transcriptomic changes we performed a meta-analysis comparing all 323 ALS versus 106 control iPSNs, accounting for batch effects between datasets (see Methods). We found 43 differentially expressed genes in pan-ALS versus control iPSNs, with 20 upregulated and 23 downregulated in ALS iPSNs (FDR < 0.05, Fig. 2a, Table S3). Amongst differentially expressed genes most increased in ALS was the endoribonuclease *RNase L (RNASEL)* which regulates decay of cytoplasmic RNA and localisation of RNA binding proteins (RBPs)^29^. Using functional enrichment analysis, we found that upregulated genes in ALS were enriched in the DNA damage response (hypergeometric p = 2.2×10^−5^; *SESN1, RRM2B, TNFRSF10B*) and p53 signalling (p = 2.7×10^−5^; *CDKN1A, TP53TG3E*) whereas downregulated genes were overrepresented by DNA-binding transcription factor activity (p = 0.003; *MYOG, TBX5, POU5F1 [Oct4])* and ventral spinal cord development (p = 0.004; *LMO4, OLIG2, FOXN4*; Fig. 2b). Gene Set Enrichment Analysis (GSEA) identified significant up-regulation of the p53 signal transduction gene set (GO:0072331, n = 264) in ALS iPSNs (normalised enrichment score [NES] +1.44, enrichment P = 4.9×10^−4^; Fig. 2c).

**Fig. 2:**
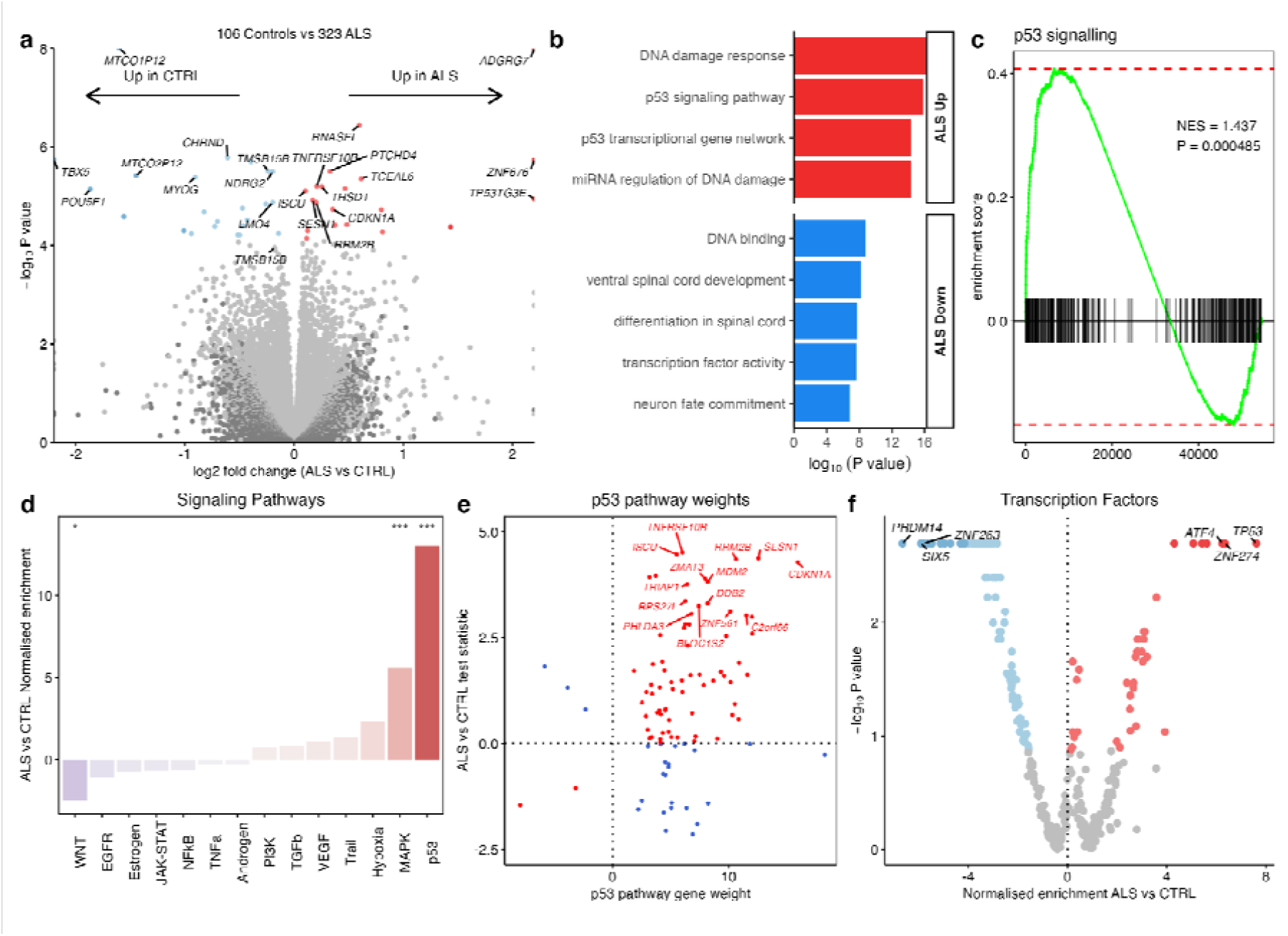
Differential gene expression in ALS versus control iPSNs. **a**, Volcano plot of differential gene expression in ALS versus control iPSNs. **b**, Functionally enriched terms in up-regulated (red) and down-regulated (blue) differentially expressed genes. **c**, Gene set enrichment analyses (GSEA) for signal transduction by p53 (GO:0072331, n = 264) in ALS versus control. NES, normalized enrichment score. **d**, PROGENy signalling pathway activities in ALS versus control. Pathways increased in ALS are red and pathways decreased are blue. *** represents P < 0.0001 and * P < 0.05. **e**, Expression changes of p53 signalling pathway genes in ALS versus control according to their PROGENy weights. Genes increasing p53 activity in ALS are red whilst genes decreasing p53 activity in ALS are blue. **f**, Activities of 429 transcription factors in DoRothEA inferred from their regulon expression changes in ALS versus control. The normalised enrichment score in ALS versus control (x-axis) is plotted according to the enrichment p-value (y-axis).

To further understand how signalling pathways are activated in ALS iPSNs, we performed a Signalling Pathway RespOnsive GENes (PROGENy)^30^ analysis which leverages perturbation experiments to more accurately infer pathway activity changes by weighting genes in each pathway based on their responsiveness. PROGENy revealed that the most substantial pathway activity increase in ALS iPSNs was in p53 (NES +13.0, p < 0.002), followed by Mitogen-Activated Protein Kinase (MAPK; NES +5.6, p < 0.002), whilst the greatest decrease was observed in WNT (NES -2.5, p = 0.03; Fig. 2d). Examining each gene in the p53 pathway according to its p53 weighting in PROGENy, revealed that the genes with the strongest responsiveness in p53 activity in ALS iPSNs included *CDKN1A, SESN1, RRM2B, MDM2, C2orf66, ZNF561* and *ZMAT3* (Fig. 2e). We next inferred the activities of 429 transcription factors (TFs) from their regulon expression within the DoRothEA database^30^. Remarkably, this revealed that TP53 was the TF with the greatest increase in activity in ALS (NES +7.62, p < 0.002) followed by ZNF274 and ATF4. The strongest TF decreases in ALS were in PRDM14, ZNF263, and SIX5 (Fig. 2f; Table S4). Interrogating individual genes constituting the TP53 TF regulon revealed the greatest increases in ALS iPSNs in *TNFRSF10B, SESN1, RRM2B, CDKN1A, ZMAT3* and *MDM2*.

Although our statistical design adjusts for dataset batch effects, it is plausible that changes between ALS and control groups were confounded by imbalances between total Ribo-Zero and poly(A) libraries. To address this, we performed a subgroup analysis in poly(A) datasets (10 datasets; 48 ALS, 43 control iPSNs) and total Ribo-Zero (5 datasets; 275 ALS, 63 control iPSNs) separately (Extended Data Fig. 8a-b). Comparing ALS versus control iPSNs revealed that in poly(A) datasets there were 69, and in total Ribo-Zero datasets there were 12 differentially expressed genes (Extended Data Fig. 8c-d). Overlapping differentially expressed genes between analyses revealed that RNase L was significantly increased in ALS iPSNs in both library preparation analyses independently. Furthermore, we confirmed significant increases in p53 pathway activity in ALS in both library preparation analyses (polyA datasets: NES +11.4, p < 0.002; Ribo-Zero datasets: NES +7.3, p < 0.002; Extended Data Fig. 10e-f). Likewise, TP53 TF activity was significantly increased in ALS iPSNs in both library preparations (polyA datasets: NES +6.54, p < 0.002; Ribo-Zero datasets: NES +5.12, p < 0.002; Extended Data Fig. 8g-h). This indicates that library preparation was not responsible for the DNA damage response gene expression changes observed in ALS iPSNs.

### p53 activation is common across ALS genetic backgrounds

To identify how ALS iPSN changes compare between ALS genetic backgrounds, we next examined the effect of each genetic subgroup on gene expression separately. 2 sALS datasets (208 sALS vs 56 controls), 7 C9orf72 datasets (60 C9orf72, 83 controls), 5 SOD1 datasets (20 SOD1 vs 63 controls), 5 FUS datasets (14 FUS vs 55 controls) and 3 TARDBP datasets (10 TARDBP vs 48 controls). Controls from each dataset were utilised only if the dataset had samples from the relevant genetic background. Although we found large numbers of differentially expressed genes (adjusted P < 0.05) in *TARDBP* mutants (3,547), the other subgroups showed more modest changes: *FUS* (239), *C9orf72* (161), and *SOD1* (7). Despite sALS being the most well-powered, with 208 iPSNs, there were only 4 differentially expressed genes (Fig. 3a-e). Correlation of transcriptome-wide gene expression changes between ALS genetic backgrounds revealed weak associations, with the strongest correlation between *SOD1* and sALS lines (Pearson R = +0.38) and the weakest between *SOD1* and *TARDBP* (R = -0.14; Fig. 3f, Extended Data Fig. 9a). To identify whether different ALS genetic backgrounds exhibit differential expression within the same genes, we overlapped genes significantly changed in expression (Table S5). Although no genes were significantly changed in expression across all ALS genetic backgrounds, Uroplakin *UPK3BL1* and nuclear pore complex interacting protein *NPIPA8* were changed in the *C9orf72, TARDBP* and *FUS* subgroups (Extended Data Fig. 9b). Functional enrichment analysis of differentially expressed genes in each genetic subgroup revealed that *C9orf72* mutants upregulated genes involved with p53 and the DNA damage response whilst downregulating cytoskeleton and microtubule genes. *FUS* mutants showed upregulation of genes involved with transcription and DNA-binding and downregulation of synaptic signalling genes. Conversely, *TARDBP* mutants upregulated neuronal and synaptic genes and downregulated genes involved in the cell cycle and RNA splicing (Extended Data Fig. 9c-e). There were no functional terms enriched amongst SOD1 or sALS differentially expressed genes.

**Fig. 3:**
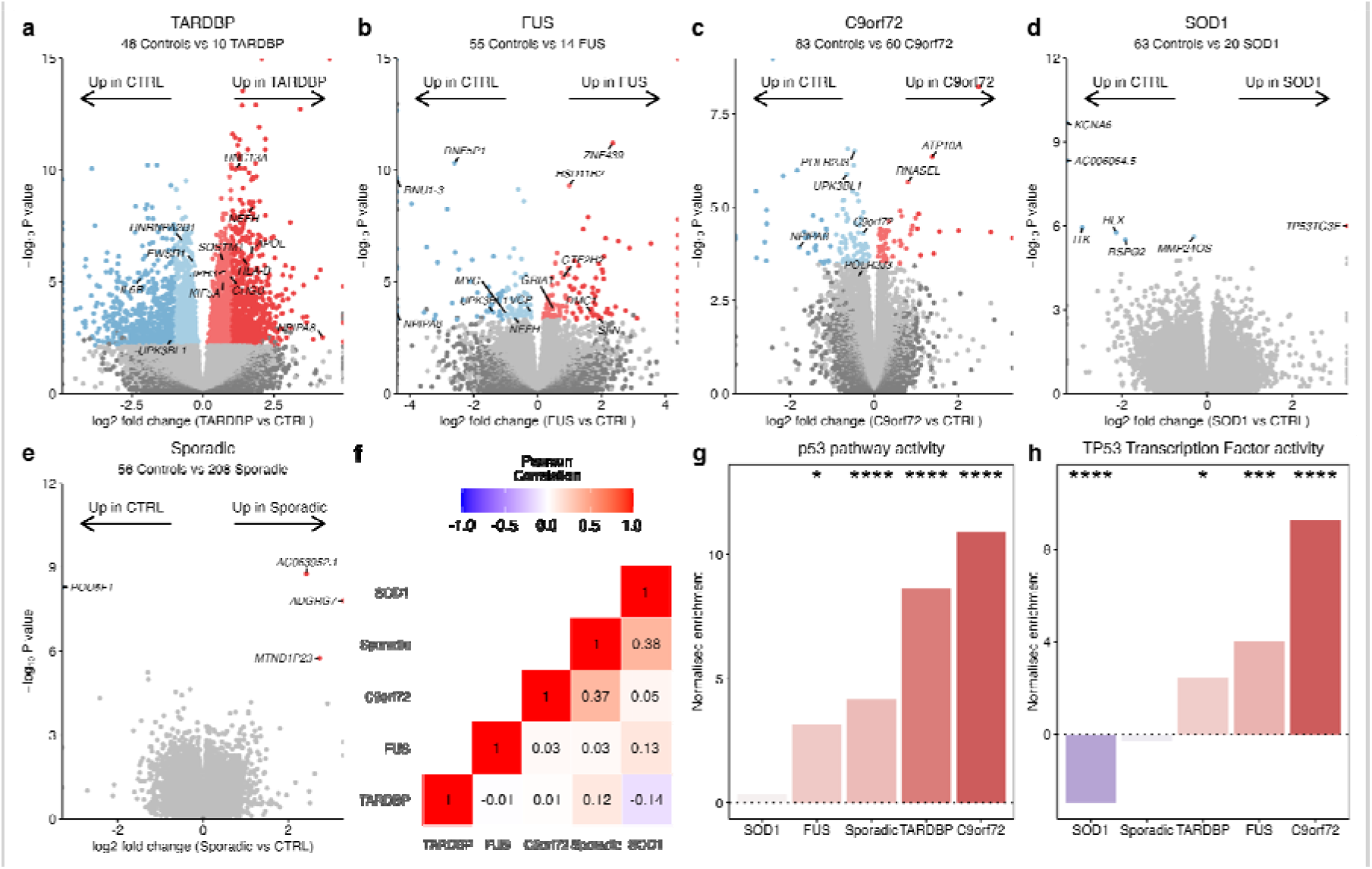
Gene expression changes in each ALS genetic background. **a-e**, Volcano plots comparing ALS iPSNs to controls in each ALS genetic background. Genes coloured red are significantly increased in the ALS subgroup and genes coloured blue are decreased in the ALS subgroup. **f**, Heatmap showing the Pearson’s correlation coefficient for transcriptome-wide changes between each genetic background. **g**, PROGENy p53 signalling pathway activity amongst each of the genetic backgrounds independently. **h**, Dorothea TP53 transcription factor regulon activity in each genetic background. **** represents P < 0.0001, *** P < 0.001, ** P < 0.01, * P < 0.05.

Examining PROGENy pathway activities in each genetic subgroup independently revealed that apart from *SOD1* (NES +0.33, p = 0.16), the p53 pathway activity was significantly increased in each of *C9orf72* (NES + 10.9, p < 0.002), *TARDBP* (NES +8.6, p < 0.002), sALS (NES +4.2, p < 0.002) and *FUS* (NES +3.2, p = 0.018; Fig. 3g). Examining the other signalling pathways revealed increases across most genetic backgrounds in hypoxia, VEGF and MAPK and decreases in WNT and PI3K (Extended Data Fig. 10a). Examining TF regulon activity in each genetic background separately revealed significantly increased TP53 activity in *C9orf72* (NES +9.3, p < 0.002), *FUS* (NES +4.0, p = 0.004) and *TARDBP* (NES +2.5, p = 0.01) but decreased activity in *SOD1* (NES -3.0, p < 0.002) whilst *sALS* (NES - 0.31, p = 0.25) was non-significantly changed (Fig. 3h). Observing the other TF activity changes between genetic backgrounds revealed 5 TFs that were significantly changed in the same direction across 4 of 5 genetic backgrounds, of which ZNF274 was increased whilst GATA3, MAZ, TAL1 and TEAD4 were decreased in activity in the ALS subgroups (Extended Data Fig. 10b). Taken together, despite transcriptome-wide heterogeneity between genetic backgrounds, these data suggest that p53 signalling activation is common across the ALS spectrum in iPSNs.

### ALS postmortem tissue shows p53 activation

To identify whether iPSN ALS gene expression signatures are also found in postmortem tissue, we compared our findings with postmortem spinal cord RNA-seq from the NYGC ALS cohort, consisting of tissue from 153 ALS patients and 80 controls^5,31^. We found 14,529 differentially expressed genes in postmortem ALS versus control spinal cord samples, with 6,417 upregulated and 8,112 downregulated in ALS (FDR < 0.05; Extended Data Fig. 11a). *CHIT1, GPNMB* and *LYZ* were the most strongly upregulated genes in ALS spinal cord, consistent with a recent report^5^. Functional enrichment analysis revealed that upregulated genes were enriched in the stress response, programmed cell death and the DNA damage response, whilst downregulated genes were enriched in neuronal functions (Extended Data Fig. 11b). As with iPSNs, signalling pathway and TF regulon analysis confirmed that ALS postmortem spinal cord was significantly upregulated in both p53 signalling (NES +4.7, p < 0.002) and TP53 activity (NES +4.01, p = 0.05; Extended Data Fig. 11c-d). Other signalling pathways also significantly increased in ALS were TNFα, Androgen, NFκB and hypoxia whereas EGFR and VEGF pathways were significantly decreased in activity.

Correlating transcriptome-wide ALS gene expression changes between iPSNs with postmortem spinal cord revealed a weak positive correlation (Pearson R = +0.13; Extended Data Fig. 11e). Of the 43 differentially expressed genes changed in ALS iPSNs, 16 (37.2%) were also changed in ALS postmortem spinal cord, with 7 co-upregulated and 9 co-downregulated in both ALS iPSNs and postmortem (hypergeometric test p = 0.0002; Table S6). Amongst the co-upregulated genes were the DNA damage response and p53 pathway genes *CDKN1A, TNFRSF10B, SESN1* and *RRM2B* as well as the endoribonuclease *RNase L (RNASEL)* and oxidative stress responder *ISCU*. These results not only confirm p53-dependent DNA damage response upregulation in ALS but the overlapping genes between iPSNs and postmortem offer insight into motor neuron-specific changes across ALS that start early and persist into the later stages of the disease.

Examining differential expression in each postmortem spinal cord genetic subgroup revealed increases in both p53 signalling and TP53 activity in each of sALS (n = 115; p53: NES +5.0, p < 0.002; TP53 NES +4.1, p = 0.03), *C9orf72* (n = 29; NES +4.7, p < 0.002; TP53 NES +4.4, p = 0.16), *SOD1* (n = 5; NES +3.4, p = 0.07; TP53 NES +2.1, p = 0.1) and *FUS* (n = 2; NES +2.6, p = 0.6; TP53 NES +3.1, p = 0.18; Extended Data Fig. 11f-g). Correlating transcriptome-wide ALS changes in postmortem spinal cord with iPSNs for each genetic subgroup revealed weak associations with the strongest between C9orf72 (Pearson R = +0.25) followed by sALS (R = +0.15), SOD1 (R = +0.07) and FUS (R = +0.002). C9orf72 mutant iPSNs and postmortem spinal cord shared 20 differentially expressed genes including axon dynein complex genes DNAH17, DNAAF1, and TEKT2, whereas FUS shared 6 and sALS shared 1 overlapping differentially expressed gene (Extended Data Fig. 11h).

### Splicing alterations in RBPs and neuronal genes

A proposed molecular mechanism of disease in ALS motor neurons is dysregulated alternative splicing, which may contribute to genomic instability underlying DNA damage response activation^32,33^. To identify alternative splicing changes in ALS iPSNs, we utilised the splice graph tool MAJIQ^34,35^, which quantifies local splicing variations from large heterogeneous RNA-seq datasets and corrects for dataset batch effects (Fig. 4a)^36^. Since total Ribo-Zero RNA libraries predominantly capture unprocessed nascent pre-mRNAs and poly(A) selected RNA libraries capture mature poly-adenylated mRNAs, we restricted splicing analyses to iPSNs that had undergone poly(A) library preparation (10 datasets composed of 48 ALS and 43 control iPSNs). Comparing ALS versus control iPSNs identified 264 local splice variation events in 161 unique genes that were significantly different between ALS and control (TNOM p < 0.05, Δ PSI > 0.1; Fig. 4b, Table S7). Amongst the splicing events most changed in ALS were in genes that are involved in DNA repair (including *POLM, METTL22, HUWE1, HDAC1, MTA1, PMS1, ZSWIM7*) and RBPs (including *YTHDC2, THOC1, PRR3, STAU2, PTBP3, SREK1, POLDIP3*; Fig. 4c). The event in *POLDIP3* was the same exon skipping event that occurs upon TDP-43 depletion (Extended Data Fig. 12a-b)^37^, indicating that TDP-43 nuclear loss of function may contribute to splicing changes in ALS iPSNs. Functional enrichment analysis of the 161 genes containing differential splicing showed enrichment in protein binding, synaptic and neuronal functions (Fig. 4d), which are central to ALS motor neuron physiology.

**Fig. 4:**
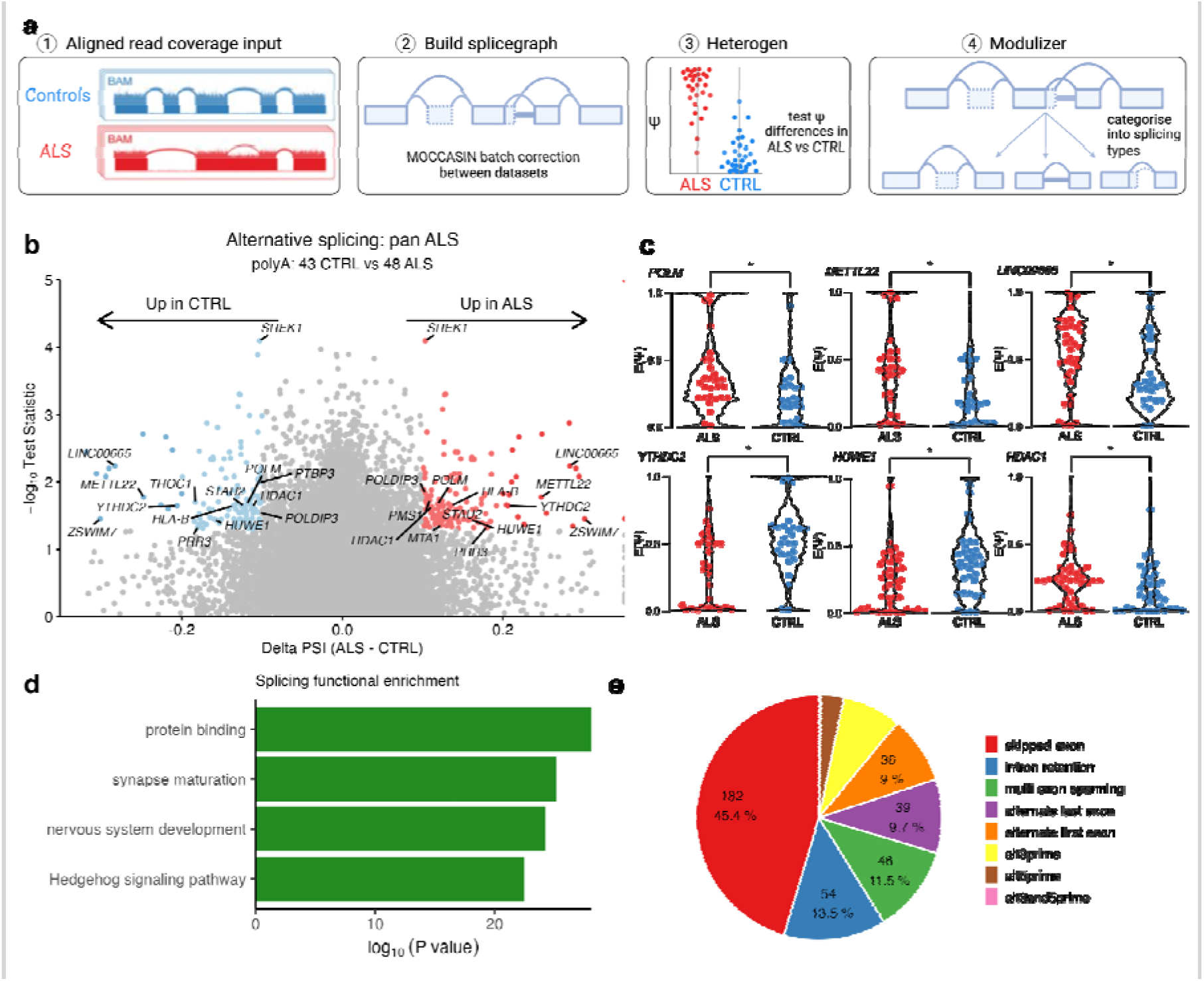
Alternative splicing alterations in ALS iPSNs. **a**, Schematic of the steps in splicing analysis of ALS and control iPSNs with MAJIQ^35^. **b**, Differential alternative splicing in poly(A) selected ALS versus control iPSNs. Y-axis is -log_10_ of TNOM p-value with events < 0.05 coloured. X-axis is Δ PSI (ALS - CTRL) with events > 0.1 coloured red (increased) and < -0.1 blue (decreased). **c**, Violin plots showing PSI values (y-axis) for each ALS (red) and control iPSN samples (blue) for the representative splicing events in *POLM, METTL22, LINC00665, YTHDC2, HUWE1 and HDAC1*. **d**, Functionally enriched terms amongst genes with differential alternative splicing. **e**, Pie chart showing the categorisation of differential local splice variants into each of the basic splicing event types using the <u>MAJIQ modulizer</u>.

The local splice variations identified by MAJIQ are predominantly complex, being composed of combinations of various 3’ and 5’ splice sites rather than simple binary events (e.g. exon skipping or intron retention [IR]). Furthermore, splice events are not restricted to annotated reference transcriptome splice sites and of the 264 differential splicing events, 28 (10.6%) involved de novo splice junctions. Of these, 7 were found to be de novo (cryptic) exons (*RELCH, HOXC4, RBM26, SLC35B3, TENM3, TPTEP2-CSNK1E, ZSCAN29*) although none of these overlapped with TDP-43 depletion^37^. 50 out of 264 (18.9%) differential splice events harboured IR within the local splice variation. Breaking down each local splice variation into its component splice types and categorising these into basic splicing modules revealed that exon skipping was the most common splicing type (182, 45.4%) followed by IR (54, 13.5%; Fig 4e). Splicing alterations, particularly IR, result in cotranscriptional RNA:DNA hybrids (R-loops) that predispose to genomic instability and DNA double-strand breaks^38,39^. We propose that splicing defects and R-loop formation in ALS iPSNs may contribute to genome instability and induce the DNA damage response. In support of this, mutations in the RNA:DNA helicase senataxin (*SETX*), which usually resolves R-loops, cause young-onset ALS^40^ and depletion of FUS and TDP-43 both lead to R-loop associated DNA damage^41^.

### ALS iPSNs are enriched in SNVs, indels and gene fusions

Genome instability triggers the DNA damage response and p53 signalling^42^. To explore the possibility that DNA damage arises in ALS iPSNs we ran the GATK variant discovery pipeline, which detects single nucleotide variants (SNVs), insertions and deletions (indels), that are signatures of aberrant repair of DNA damage. Variant detection is highly sensitive to coverage and sequencing chemistries and so we restricted variant detection to the Answer ALS dataset (ALS n = 238, CTRL n = 42 iPSNs). To increase the likelihood that identified variants were DNA variants we excluded known RNA editing sites. Across all filtered variant types, we found significantly greater numbers of variants per iPSN in ALS compared to control (Wald test p < 2×10^−16^; Fig. 5a). Examining each variant type separately, revealed significantly greater numbers per iPSN of SNVs (Wald test p < 2×10^−16^), insertions (p = 2.5×10^−14^) and deletions (p = 1.1×10^−9^) in ALS compared to control (Fig. 5b; Table S8). We observed the greatest increase in the number of C>T base substitutions in ALS iPSNs relative to controls (p < 2×10^−16^), followed by T>A (p = 0.001), C>T (p = 0.001) and C>A (p = 0.004; Fig 5c). By observing the relative proportions of each type of base substitution, we found ALS iPSNs showed increases in T>C, T>A and C>A whereas the proportions of C>T, C>G and T>G were decreased relative to controls (Extended Data Fig 13a). Examining the number of variants per iPSN in each ALS genetic background revealed significantly greater numbers of variants only in the sALS subgroup, although the other mutant subgroups were relatively underpowered when restricted to Answer ALS (*sALS* n = 200, *C9orf72* n = 21, *SOD1* n = 8, *TARDBP* n = 1, *FUS* n = 0; Extended Data Fig. 13b).

**Fig. 5:**
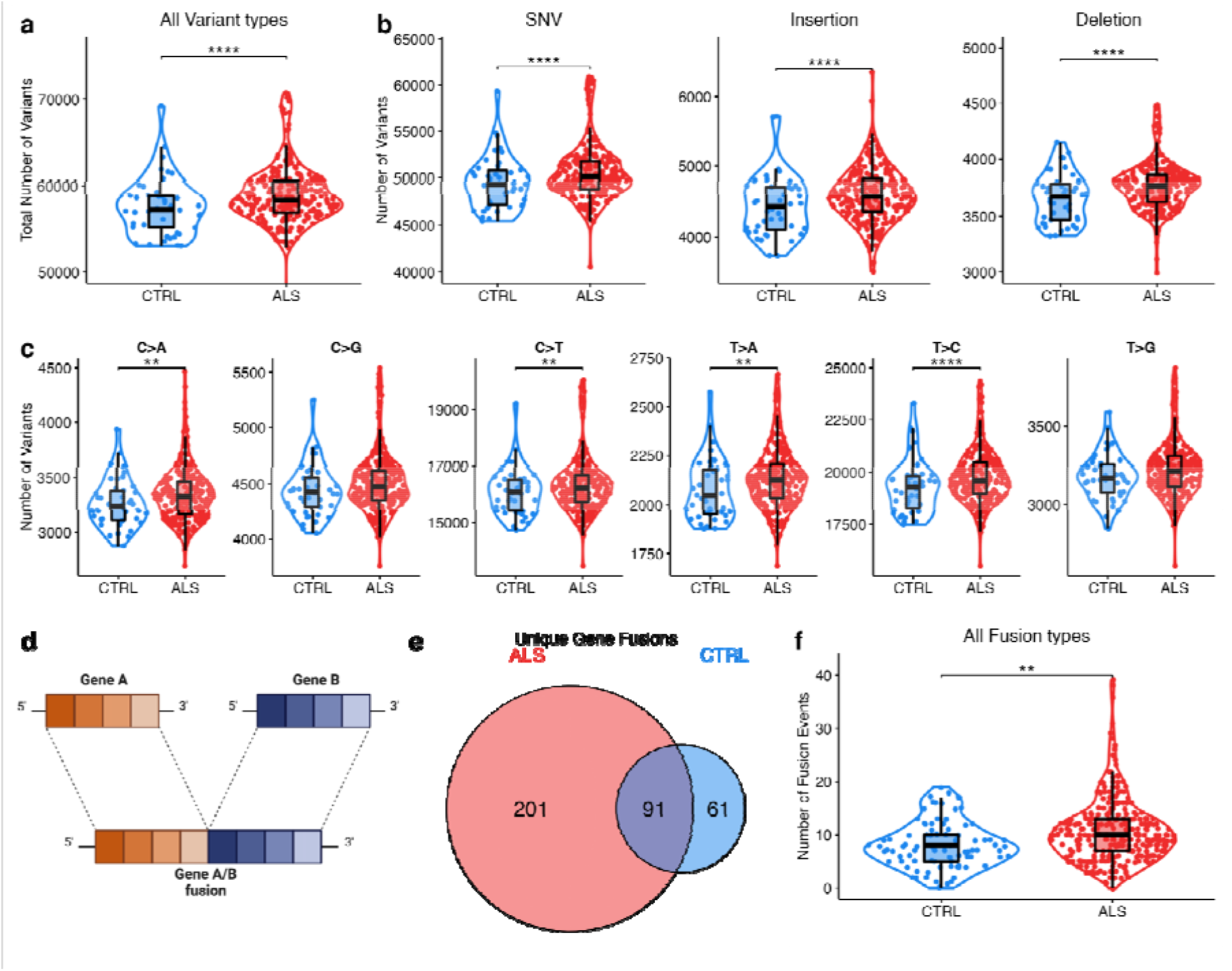
ALS iPSNs exhibit greater numbers of SNVs, indels and gene fusions. **a**, Violin plot showing the numbers of variants identified per iPSN in ALS (red) and CTRL (blue) samples from Answer ALS. **b**, Violin plots showing the number of variants according to their annotated type (Single Nucleotide Variant [SNV], Insertions, Deletions). **c**, Number of SNVs classified according to their base substitution type in ALS and CTRL. **d**, Schematic of a gene fusion. **e**, Venn diagram overlapping unique gene fusions in ALS and CTRL iPSNs from paired-end datasets. **f**, Violin plot showing the numbers of unique gene fusions per iPSN. **** represents P < 0.0001, *** P < 0.001, ** P < 0.01, * P < 0.05.

Gene fusions are another important class of genome alteration that can arise from the repair of DNA damage. Fusions involve two genes becoming juxtaposed due to structural rearrangements, including inversions and translocations (Fig. 5d). To explore the possibility of whether ALS iPSNs exhibit increased numbers of gene fusions we examined for such events using the RNA-seq iPSN datasets. Accurate gene fusion detection requires paired-end reads and so we restricted the STAR Fusion analysis to the 11 paired-end datasets (306 ALS, 90 CTRL iPSNs, Table 1)^43^. Since gene fusion discovery is also sensitive to differences in read coverage, we adjusted for the total read count per sample as well as dataset batch effects in the generalised linear model. There were a total of 292 unique gene fusions observed in ALS iPSNs and 152 unique gene fusions in control iPSNs, with 91 shared in both conditions (Fig. 5e). Amongst the 201 gene fusions identified in ALS iPSNs but not in controls, were fusions affecting genes implicated in ALS including VAPB–APCDD1L-DT, ATXN1–ZFYVE27, TUBA1A–NEFM and OSTF1–APP (Table S9). Furthermore, of the 292 genes fusions identified in ALS iPSNs, 14 affected genes also exhibited altered splicing, supporting the possibility of *trans*-splicing, that post-transcriptionally joins exons from separate pre-mRNAs^44^. By comparing the proportion of each unique gene fusion in ALS with CTRLs iPSNs, we identified 9 gene fusions with a significantly greater burden in ALS iPSNs (Table S9). Interestingly, these mostly involved long noncoding RNAs (lncRNAs), for example, the gene fusion with the greatest burden in ALS was a neighbour fusion between the lncRNA LINC01572 and PMFBP1 (OR 3.3, 95% CI 1.6-Inf, Fisher’s exact test p 0.001).

Examining the frequency of gene fusions per iPSN, revealed significantly greater numbers of gene fusions in ALS compared to controls (Wald test p = 0.002; Fig. 5f; Table S10). Comparing the frequencies of each type of gene fusion between ALS with control iPSNs, revealed trends towards increased numbers in ALS iPSNs for each of gene neighbours (Wald test p = 0.13), overlapping neighbours (p = 0.83), local inversions (p = 0.95), distant intra-chromosomal (p = 0.93), although this was statistically significant only for inter-chromosomal fusions (p = 0.0002; Extended Data Fig. 13c). Comparing the number of gene fusions per iPSN in each ALS genetic group with their respective dataset controls, revealed significantly greater numbers of gene fusions in C9orf72 repeat expansions (Wald p = 7×10^−5^) and SOD1 mutant groups (Wald test p = 6.6×10^−5^; Extended Data Fig. 13d). Taken together, these findings reveal enrichment of SNVs, indels and gene fusions in ALS motor neurons, which we propose is a genomic signature arising from elevated DNA damage.

## Discussion

Here we present a comprehensive catalogue of ALS motor neuron transcriptomes, comprising >400 iPSNs spanning 10 different ALS mutations and sALS. Systematically integrating this data provides substantially improved statistical power to detect perturbations in ALS motor neurons. Our major finding is that ALS motor neurons display augmented DNA damage response and p53 signalling activation across the spectrum of ALS. ALS motor neurons showed altered splicing in neuronal genes as well as greater numbers of SNVs, indels and gene fusions compared to controls, which may contribute to the elevated DNA damage response. Our findings add to a growing body of evidence for the role of defective DNA repair and induction of the DNA damage response in ALS motor neurons^45^.

Despite the large sample size, we found relatively few differentially expressed genes across the ALS spectrum. This is likely due to heterogeneity between ALS genetic backgrounds since these displayed weak transcriptome-wide correlations, thus limiting the detection of common gene expression changes at the pan-ALS level. Whilst the sALS subgroup was the most well powered, representing two-thirds of all ALS iPSNs, it showed only 4 differentially expressed genes, reminiscent of what we found in sALS iPSC-derived astrocytes^46^. sALS represents a genetically heterogeneous group and likely includes patients carrying as yet unknown pathogenic gene mutations^47^. Additionally, environmental risk factors play an important role in ALS aetiology particularly in patients without a highly penetrant mutation^48–52^. Whilst the iPSC model is an elegant approach to model sALS, a notable limitation is that it does not reproduce the patients’ environmental exposures. Nonetheless, iPSNs without a known ALS mutation still showed significantly elevated p53 activity.

In summary, we integrated all publicly available transcriptomic data of iPSNs from ALS donors, to enable a greater understanding of the determinants of motor neuron dysfunction. Future studies are required to identify whether DNA damage in ALS motor neurons results from increased spontaneous damage or impaired repair. Identifying the identity and loci of the DNA lesions that accumulate in ALS motor neurons driving p53 activation will help in this regard and may expose desperately needed tangible therapeutic targets in this devastating and currently untreatable disease.

## Data Availability

All raw and processed sequencing data generated in this study are available on public repositories under accession numbers shown in Table 1.

## Methods

### Eligibility criteria and search strategy

We evaluated all human iPSN datasets that had undergone bulk RNA-sequencing (RNA-seq) that examined ALS and controls (healthy individuals or isogenic correction), regardless of the iPSN differentiation protocol or RNA-seq library strategy. All ALS subtypes were included and the definition of ALS used by each individual dataset was accepted. In datasets with multiple timepoints through motor neuron differentiation only the final most terminal differentiated timepoint was utilised^13^. We excluded datasets that had not undergone a motor neuron differentiation protocol or failed iPSN identity or RNA-seq quality control measures. We systematically reviewed RNA-seq databases, including Gene Expression Omnibus (GEO), NCBI sequence read archive (SRA), EBI arrayExpress, European Nucleotide Archive (ENA), synapse.org and manually searched reference lists as well as ALS data portals of relevant studies up to June 2022. The search strategy included keywords relating to ALS and motor neurons.

### RNA-seq processing, integration and quality control

The iPSN differentiation protocol method (including induction, specification and terminal differentiation) as well as RNA-seq library strategy (RNA extraction, library preparation, sequencing instrument and read metrics) for each dataset is noted in Table S1. Raw RNA-seq reads (fastq files) and accompanying metadata were downloaded using nfcore/fetchngs v1.5 pipeline ^53^ and pysradb v1.3 using the sample SRA accession number. Reads were processed using the nfcore/rnaseq v3.8.1 pipeline^53^. Raw reads underwent adaptor trimming with Trim Galore, removal of ribosomal RNA with SortMeRNA, alignment to Ensembl GRCh38.99 human reference genome using splice-aware aligner, STAR v2.7.1 and BAM-level quantification with Salmon. Samples were subjected to extensive RNA-seq quality control utilising FastQC, RSeQC, Qualimap, dupRadar, Preseq, and SAMtools and results were collated with MultiQC. Samples that passed the nfcore/rnaseq quality control status checks were included in the meta-analysis (Table S2). The median read depth was 115 (range 6 - 164) million reads per sample.

We used principal component analysis (PCA) and unsupervised clustering to interrogate the batch effects of clinical variables, iPSN protocols and RNA-seq strategies between samples and datasets. Gene counts were normalised and transformed using the variance stabilizing transformation function in DESeq2. Principal components were calculated based on the 500 highest variance genes using the plotPCA function and individual PC gene loadings were extracted with the prcomp function. Samples clustered into two groups based on library preparation (polyA or total Ribo-Zero RNA). We examined the motor neuron transcriptomic identities of iPSNs by clustering using the ComplexHeatmap package based on the expression of canonical neuronal and glial cell type markers as well as dorsoventral^54^ and rostrocaudal (HOX) gene markers. The Lee et al. dataset was excluded due to RNA library batch effects between ALS (Ribo-Zero) and control (polyA) samples as well as inadequate neuronal marker expression^20^. We excluded three control samples in AnswerALS that whole-genome sequencing revealed to have pathogenic ALS mutations. Additionally, 4 Answer ALS iPSNs from non-ALS motor neuron disease patients were excluded. NeuroLINCS consists of 3 distinct iPSC protocols (iMNs, diMNs and undifferentiated iPSCs; Table S1), of which only the iMN and diMN batches were included. Gender was confirmed by examining expression of the X chromosome gene *XIST* (female) and Y chromosome genes *KDM5D, DDX37, RP54Y1* and *EIFAY* (male).

### Modelling differential gene expression

STAR aligned and Salmon quantified transcript abundance were summarised at the gene-level using tximport in R v4.1.3. Differential gene expression analysis was then fitted using DESeq2^55^. The meta-analysis results of ALS iPSNs was generated by comparing the ALS versus control groups using the Wald test, controlling for sex differences and dataset variation with the design formula ∼gender + dataset + condition. This design controls for technical variation due to library preparation (nested within the dataset variable), which was the main factor driving PCA structure (Extended Data Fig. 2-3), thereby increasing the sensitivity for identifying differences due to ALS. We orthogonally estimated technical variation using the RUVg method that takes empirically defined negative control genes to estimate low-rank technical variation in the data, specifying 5 RUV factors^56^. To examine the effect of each ALS genetic background on gene expression a similar approach was used, comparing the ALS versus control samples using the design formula ∼gender + dataset + genetic_group. For these subgroup analyses, control samples from each dataset were only utilised if the dataset also exhibited the relevant ALS genetic background.

Results for each genetic background were correlated by matching the Wald test statistic for each gene followed by Pearson correlation. In all analyses, genes were considered differentially expressed at FDR < 0.05. Significantly up- and down-regulated differentially expressed genes were used as input to functional enrichment analyses, which was used to identify enriched pathways using g:Profiler2. g:Profiler2 searches the following data sources: Gene Ontology (GO; molecular functions, biological processes and cellular components), KEGG, REAC, WikiPathways, CORUM and Human phenotype ontology. In the functional enrichment bar charts, the top significant terms were manually curated by removing redundant terms. Gene Set Enrichment Analysis (GSEA) was performed using FGSEA on GO:0072331 (signal transduction by p53 class mediator) gene set. The decoupleR package was used to estimate PROGENy signalling pathway activities and DoRothEA TF regulon activities inferred from gene expression changes ^30^.

Postmortem spinal cord ALS RNA-seq samples were derived from samples from the New York Genome Centre (NYGC) ALS consortium. Samples from non-spinal cord sites were excluded as well as samples that failed quality control as described previously^5^. Processed gene counts are available from Zenodo accession <u>6385747</u>.

In cases where multiple spinal cord samples were available from donors only the cervical cord sample was included. Differential expression results for postmortem spinal cord ALS were calculated by comparing ALS versus control samples, accounting in the design for the RNA library preparation method, gender and the site of spinal cord (cervical, thoracic or lumbar).

### Alternative Splicing Analyses

All modes of alternative splicing were analysed using MAJIQ v2.4^34,35^ on poly(A) selected RNA library samples. STAR aligned BAMs were used as input to the MAJIQ splice graph builder using Ensembl GRCh38.99 transcript annotation. Batch effects between datasets and gender were corrected for by using *MOCCASIN*^*36*^. Differential splicing was calculated using the *MAJIQ heterogen* function, which is designed for examining splicing across large and heterogeneous datasets. A threshold of 10% ΔΨ and TNOM p-value < 0.05 was used to call significant splicing changes between groups. Changes in each specific class of splicing were examined using the *Voila modulize* function that breaks down the complex local splice variants into the classic binary splicing events (e.g. exon skipping or intron retention). For comparison of ALS iPSN splicing events with TDP-43 deletion, RNAseq fastq files from iNeurons were downloaded from ENA <u>PRJEB42763</u>. These were processed using nfcore/rnaseq followed by MAJIQ v2.4 using the *MAJIQ deltapsi* function, which is more appropriate for this small single batch homogenous experimental replicate dataset.

### Gene variant and fusion detection

Gene variants were detected using the nfcore/rnavar pipeline v1.0.0^53^, which is based on GATK v4.2.6 short variant discovery workflow. Variant discovery is highly sensitive to coverage and sequencing chemistries and so only the Answer ALS dataset was used for variant detection thus avoiding confounding batch effects between multiple datasets. Raw RNA-seq reads were mapped using STAR in two-pass mode. SplitNCigarReads tool was used to reformat alignments that span introns for the HaplotypeCaller. BaseRecalibrator and ApplyBQSR were used for base quality recalibration. Single nucleotide variants (SNVs) and indels were called using the HaplotypeCaller and variants were filtered using VariantFiltration specifying a minimum phred-scaled confidence threshold of 20 and minimum quality depth of 2.0. and. RNA editing variants from the REDIportal v2.0 were filtered out using VCFtools –exclude-bed module. Variants were annotated using snpEff and Ensembl VEP whilst VCFtools vcf-annotate –fill-type module was used on the filtered output to classify variants into SNVs, insertions or deletions. The characteristics of variants were assessed using the MutationalPatterns package v3.6.0, which summarises the number and proportions of each type of base substitution^57^. To compare the number of variants per iPSN in ALS versus CTRL groups, a generalised linear model was fit specifying a poisson distribution adjusting for differences in read coverage per sample using a spline: *variant_count ∼ condition + rcs(total_reads, 3)*.

Gene fusions were identified using the nfcore/rnafusion pipeline v2.0.0^53^, utilising the STAR-Fusion v1.10.1^43^ workflow on paired-end RNA-seq datasets. Raw RNA-seq reads were aligned using STAR to identify chimeric transcripts, which are defined as a part of a read aligning to one gene and another part of the same read to a different gene (split) or when each end of a paired read set aligns to different genes (spanning). STAR Fusion applies numerous filters to avoid spurious fusion detection including removing chimeric reads that overlap with sequence similar regions and removing duplicate paired-end alignments. Fusion events were filtered using the FusionFilter module default settings for spanning and split reads. Fusion events with fusion fragments per million (FFPM) < 0.1 were removed. Gene fusion events are classified using FusionAnnotator module into inter-chromosomal and intra-chromosomal. Intra-chromasomal are subclassified into local gene orientation rearrangements, neighbours (<100kb apart), overlapping neighbours (genes span overlap by at least 1 base pair) and distant (>100kb apart). To compare the number of variants or fusions per iPSN in ALS versus CTRL groups, a generalised linear model was fit specifying a poisson distribution adjusting for differences in read coverage per sample using a spline and batch effects between datasets: *fusion_count ∼ condition + dataset + rcs(total_reads, 3)*. To detect burden differences of individual gene fusions in ALS compared to CTRL iPSNs, the Fisher test was used to calculate the Odds Ratio, 95% confidence interval and p-value. In both variant and fusion analyses, to examine the effect of each ALS genetic group, controls were only utilised if the dataset also included ALS samples from the relevant genetic group.

Schematics were created with BioRender.com. All error bars in the boxplots shown represent 1.5 times the interquartile range.

## Acknowledgements

We are grateful for discussions and advice from Jesper Svejstrup, James Lee, James Briscoe, Rory Maizels, Carlos Martinez Ruiz, the MAJIQ team as well as the nfcore community, particularly Hashil Patel, Phil Ewels, Praveen Raj, Martin Proks and Annik Renevey. We thank the Francis Crick Institute scientific platforms, especially members of the high performance compute team Danny Lang and John Roche. We thank the AnswerALS and NeuroLINCS teams, particularly Terry Thompson and Barry Landin, for help with access and interpretation.

This work was funded by the Francis Crick Institute, which receives its core funding from Cancer Research UK (FC010110), the UK Medical Research Council (FC010110), and the Wellcome Trust (FC010110). O.J.Z. holds a Crick Clinical PhD Fellowship supported by the University College London Hospitals Biomedical Research Centre (BRC689/ED/CB/100130). R.P. holds an MRC Senior Clinical Fellowship (MR/S006591/1) and a Lister Research Prize Fellowship.

## Author information

### Contributions

Study was conceived and designed by O.J.Z and R.P. Gene expression, splicing and fusion analyses were performed by O.J.Z with supervisorial input from G.K., J.H., A.M.C., R.L., C.M.R., N.M. and R.P. Interpretation of differentiation protocols was performed by J.N. and interpretation of motor neuron markers was performed by J.M. Postmortem data were provided by J.H. The manuscript was written by O.J.Z. and R.P. Interpretation of data and contributions to write-up were provided by J.N., G.T., J.M., C.M.R., N.M., R.L., A.M.C., S.J.B., G.K., and J.H.

