## Extended Data for "Meta-analysis of the amyotrophic lateral sclerosis spectrum uncovers genome instability"

PRISMA flow diagram summarising search results of databases, registers, and other sources.

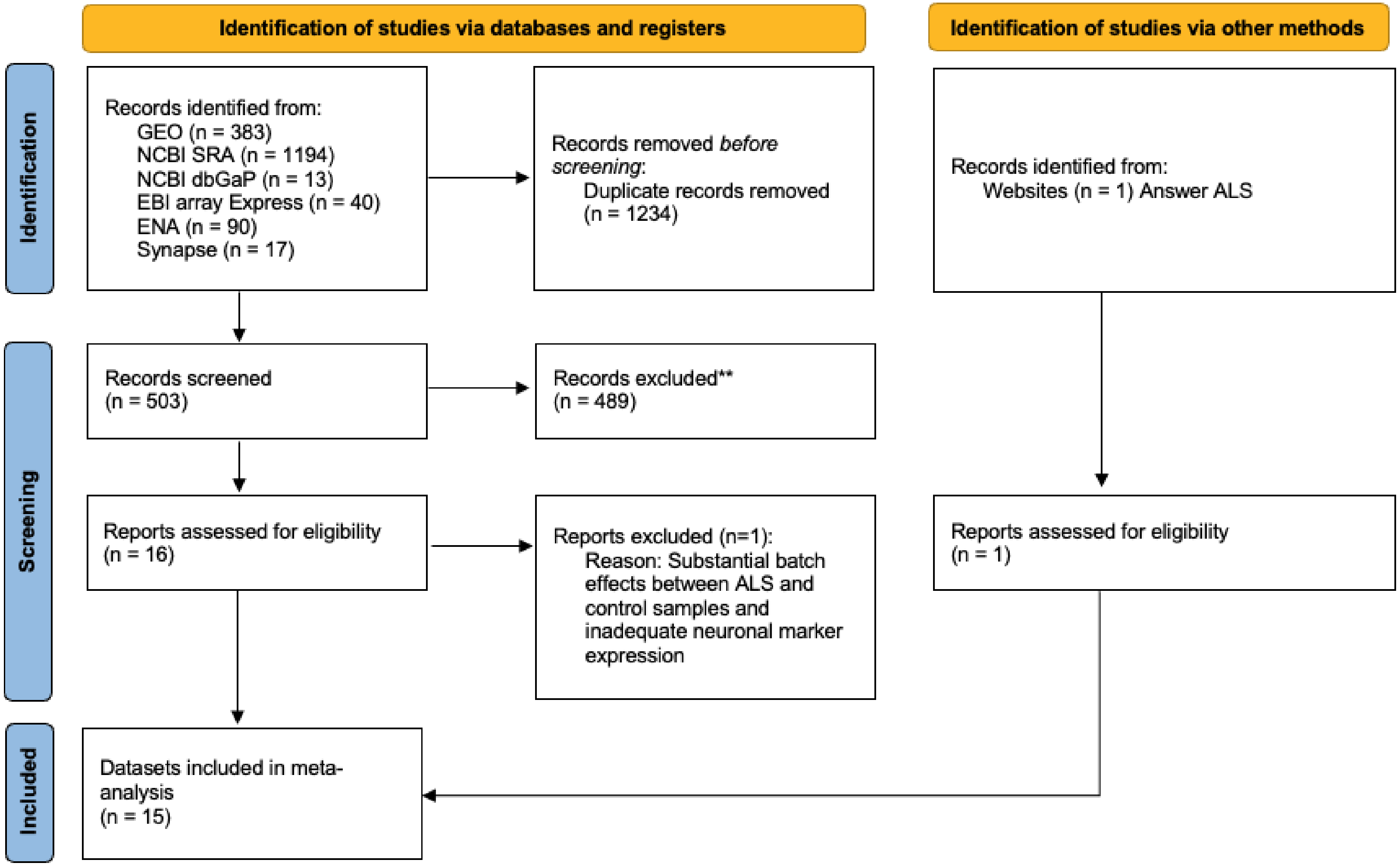


*Search updated 30th June 2022*

**Extended Data Figure 1 PRISMA flow diagram of database searches**

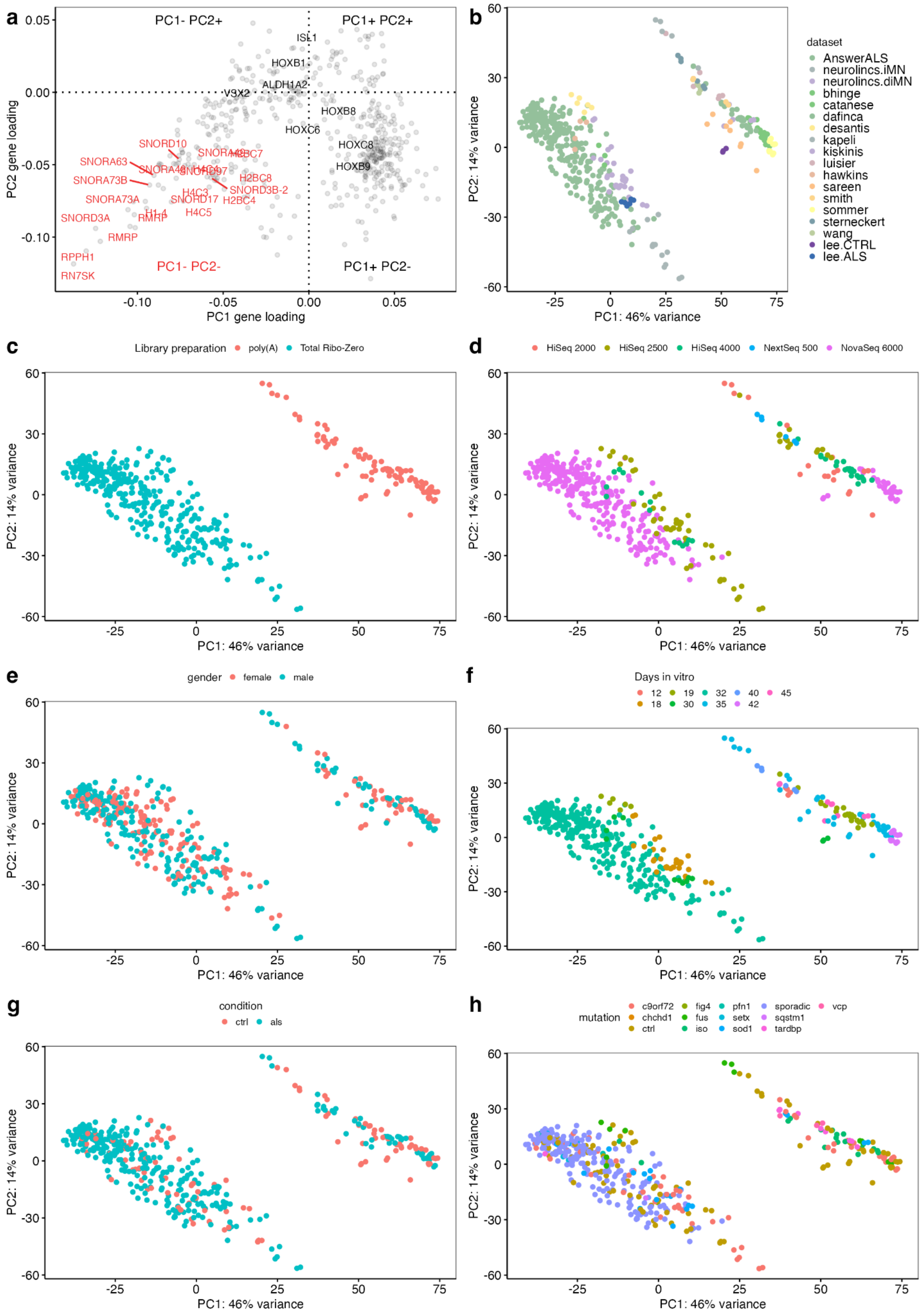

a
PC1- PC2+
PC1+ PC2+
PC1- PC2-
PC1+ PC2-
ISL1
HOXB1
ALDH1A2
V3X2
HOXB8
HOXC6
HOXC8
HOXB9
SNORD10
SNORA63
SNORA73B
SNORA73A
SNORD3A
RMRP
RPPH1
RN7SK
H1-4
H4C3
H4C5
SNORD17
H2BC4
H2BC8
SNORD3B-2
H2BC7
PC2 gene loading
PC1 gene loading
b
dataset
AnswerALS
neurolincs.iMN
neurolincs.diMN
bhinge
catanese
dafinca
desantis
kapeli
kiskinis
luisier
hawkins
sareen
smith
sommer
sterneckert
wang
lee.CTRL
lee.ALS
PC2: 14% variance
PC1: 46% variance
c
Library preparation
poly(A)
Total Ribo-Zero
d
HiSeq 2000
HiSeq 2500
HiSeq 4000
NextSeq 500
NovaSeq 6000
e
gender
female
male
f
Days in vitro
12
18
19
30
32
35
40
42
45
g
condition
ctrl
als
h
mutation
c9orf72
chchd1
ctrl
fig4
fus
iso
pfn1
setx
sod1
sporadic
sqstm1
tardbp
vcp

**Extended Data Figure 2 Principal Component Analysis coloured by sample characteristics**

a: Scatterplot of PC1 against PC2 individual gene loadings of the top 500 most variable genes. PC1- & PC2- loaded genes labelled red are histone and small non-coding RNAs, which represent non-polyadenylated transcripts. Genes labelled black are relevant to spinal motor neuron identity.

b-h: Principal component analysis of variance stabilised transformed gene expression from iPSN samples, coloured by (b) dataset, (c) RNA library preparation type, (d) sequencing instrument, (e) gender, (f) Days in vitro, (g) disease status and (h) mutation. iso, isogenic correction.

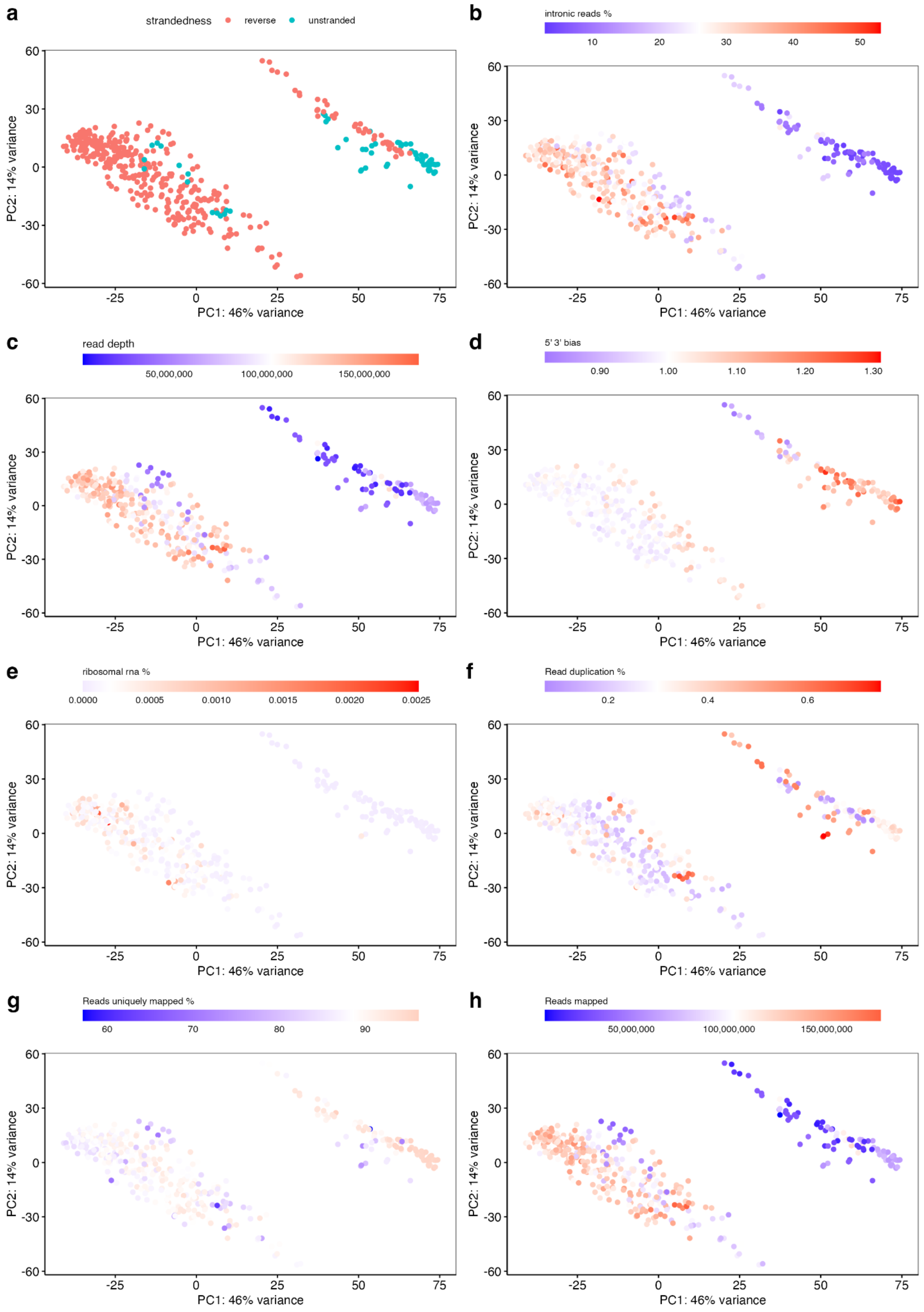

a
strandedness
reverse
unstranded
b
intronic reads %
c
read depth
d
5' 3' bias
e
ribosomal rna %
f
Read duplication %
g
Reads uniquely mapped %
h
Reads mapped
PC1: 46% variance
PC2: 14% variance

**Extended Data Figure 3 Principal Component Analysis coloured by QC metrics**

a-h: Principal component analysis of variance stabilised transformed gene expression from iPSN samples, coloured by (a) strandedness, (b) intron read %, (c) read depth, (d) Picard read duplication %, (e) ribosomal RNA % biotype contamination, (f) Qualimap 5-3' bias, (g) STAR reads uniquely mapped % and (h) Samtools raw reads mapped. Scale bars are coloured from minimum (blue) to maximum (red) value with white representing the mean value.

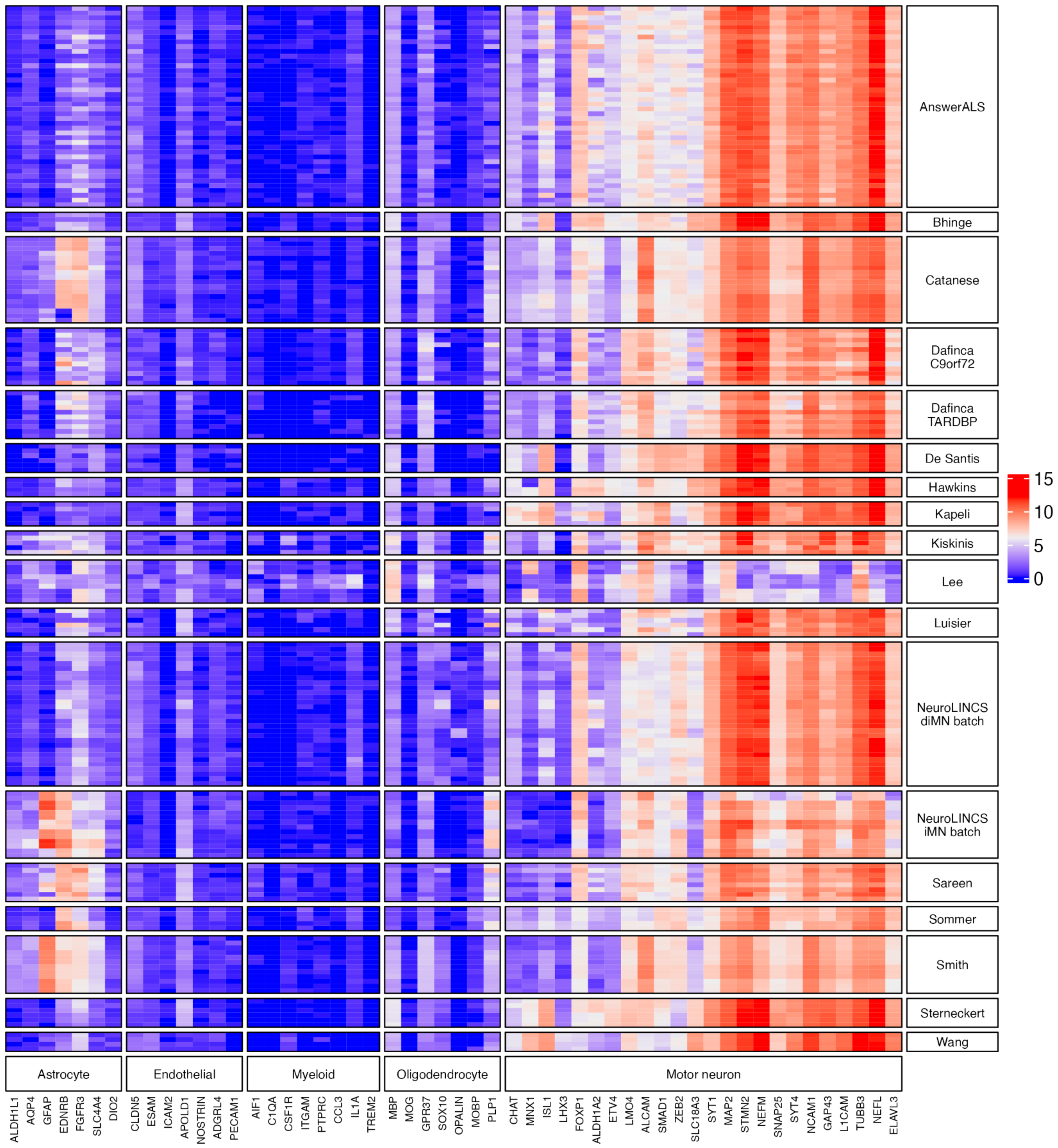


**Extended Data Figure 4 Transcriptomic identities of iPSC-derived motor neurons**

Heatmap of variance stabilised gene counts for astrocytes, endothelial cells, myeloid cells, oligodendrocytes and motor neurons (columns) across iPSC-derived motor neuron datasets (rows). To improve the visualisation of all datasets, for AnswerALS only 100 samples are plotted. Lee et al dataset is removed from the meta-analysis because of different library preparations between ALS and controls and poor neuronal marker expression.

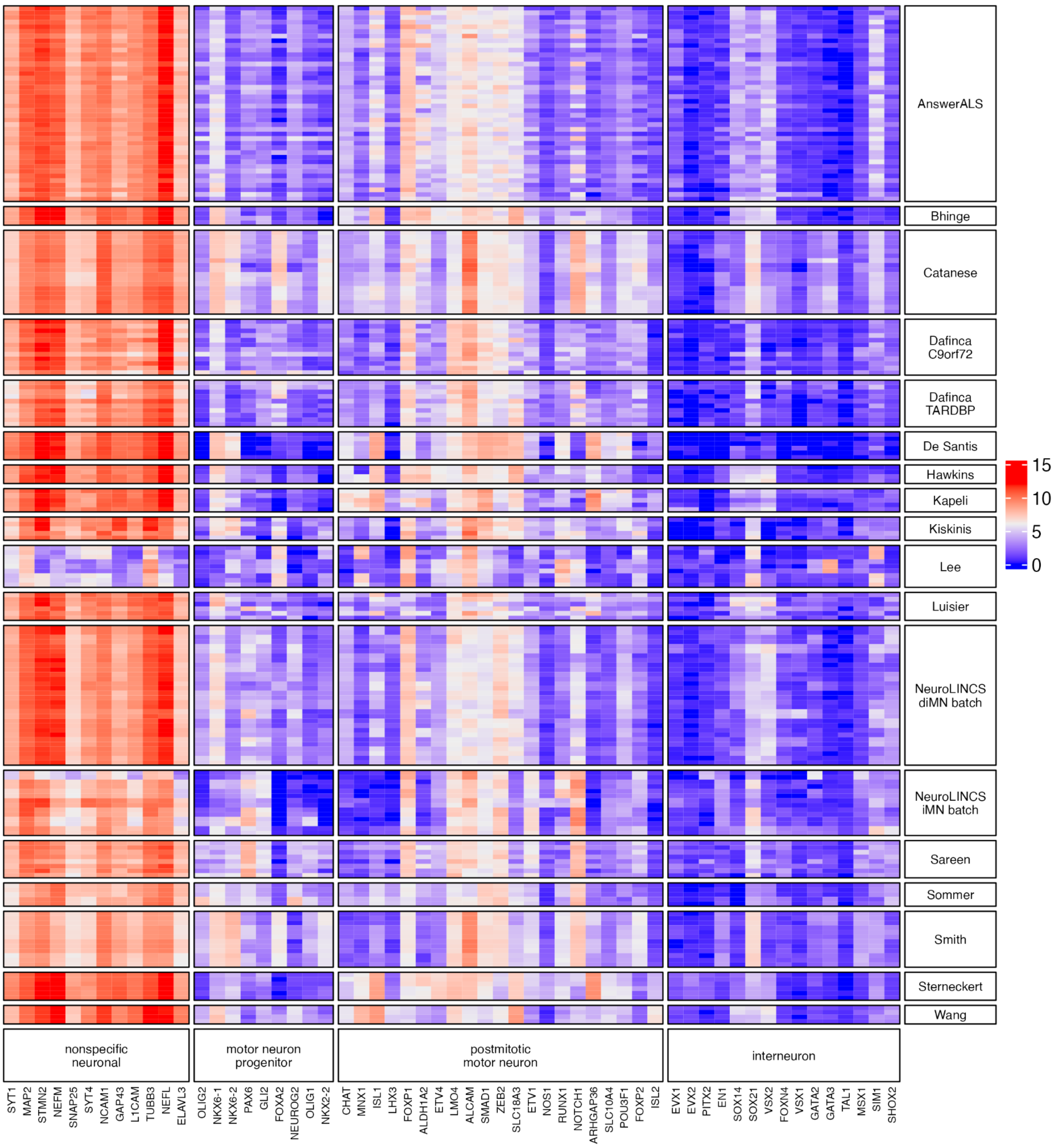


**Extended Data Figure 5 Motor neuron identities**

Heatmap depicting variance stabilised gene counts of neuronal markers (columns) across the iPSC-derived spinal motor neuron datasets (rows). To improve the visualisation of all datasets, for AnswerALS only 100 samples are plotted. Lee dataset is removed from the meta-analysis because of inadequate neuronal marker expression.

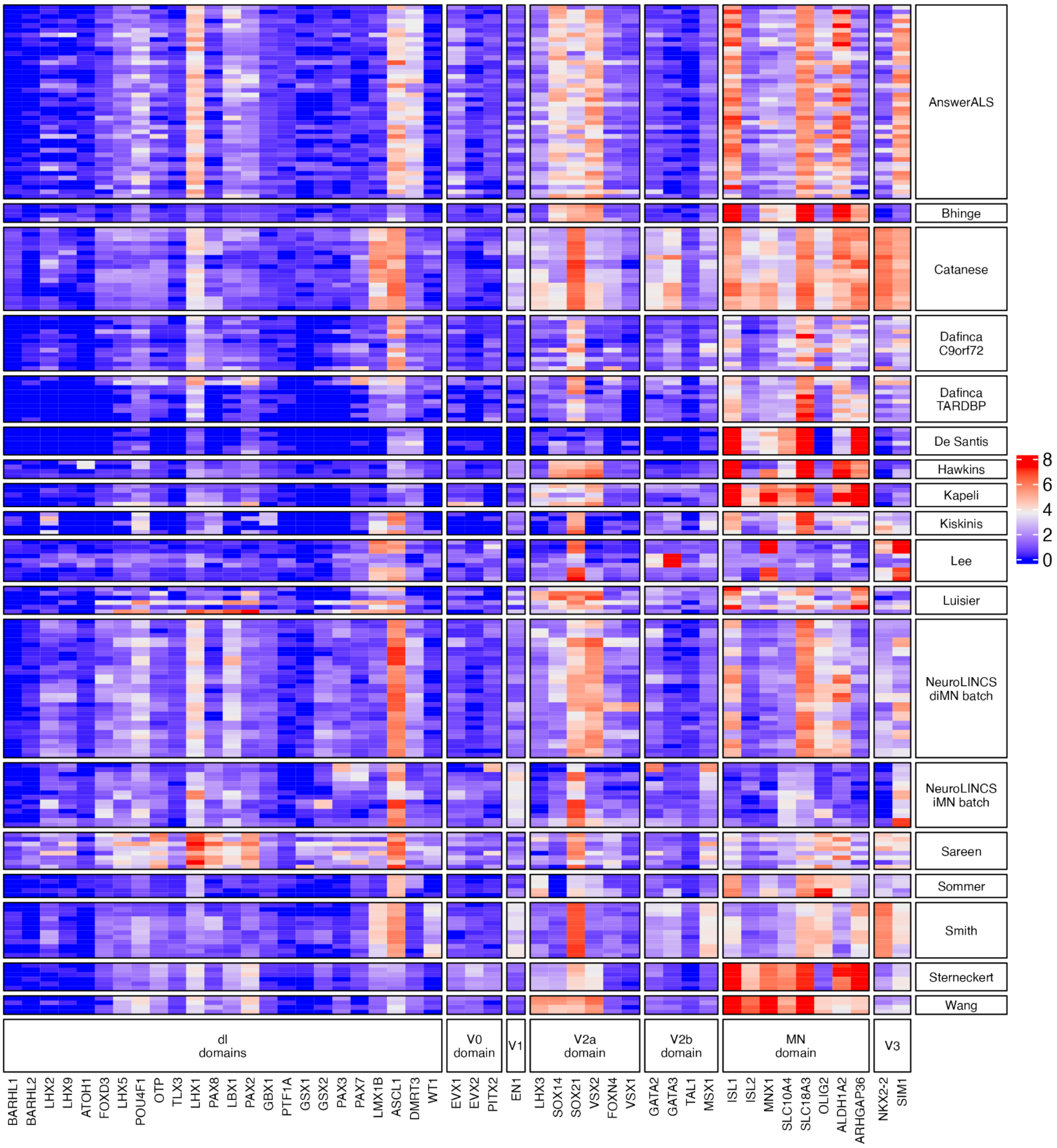


### Extended Data Figure 6 Spinal cord dorsoventral identities

Heatmap of variance stabilised gene counts for spinal cord dorsoventral markers from Raynon et al.[16] (columns) across iPSC-derived motor neuron datasets (rows). To improve the visualisation of all datasets, for AnswerALS only 100 samples are plotted.

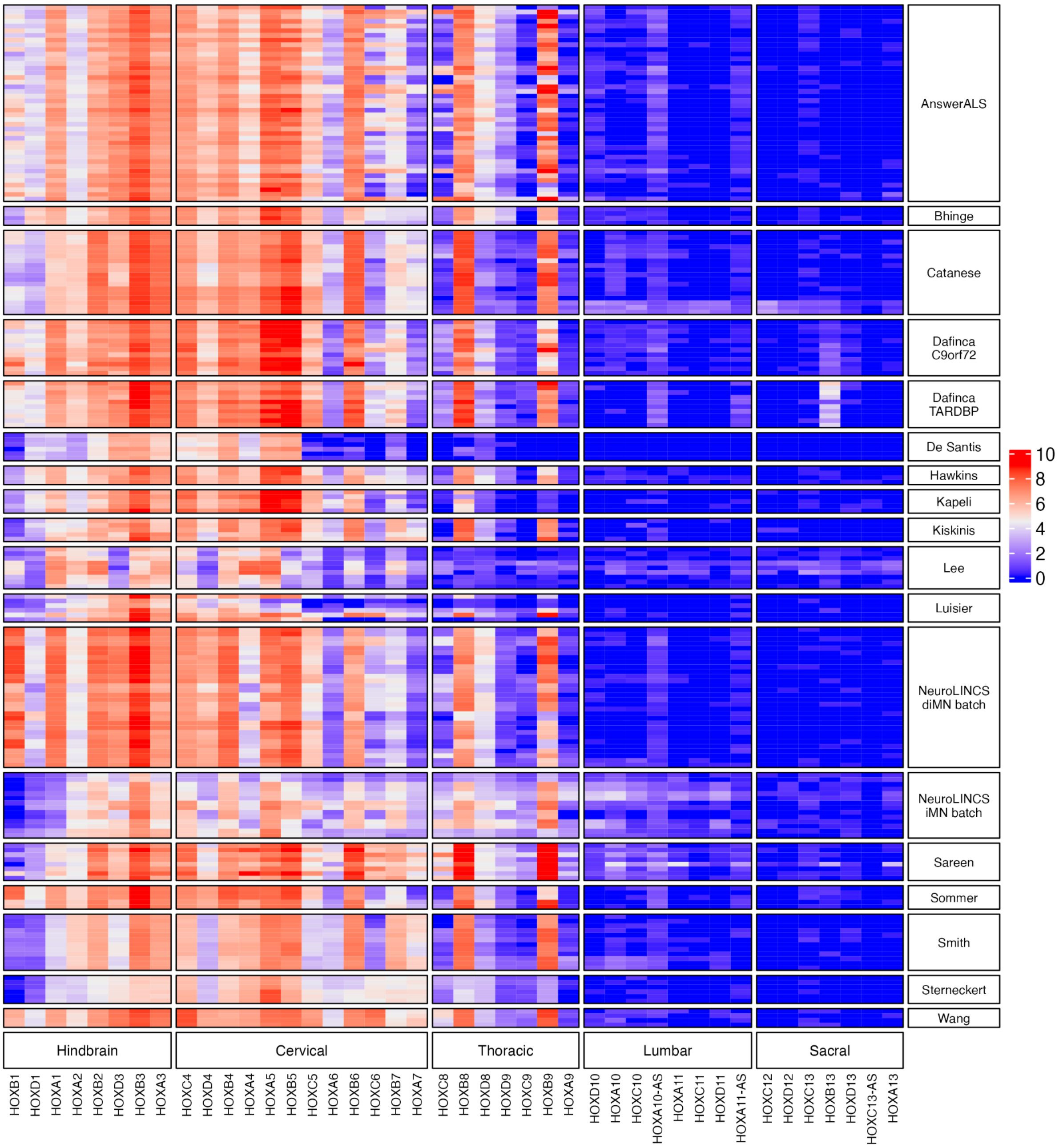


### Extended Data Figure 7 HOX marker rostrocaudal regional identities

Heatmap of variance stabilised gene counts for HOX markers (columns) across the iPSC-derived motor neuron datasets (rows). To improve the visualisation of all datasets, for AnswerALS only 100 samples are plotted.

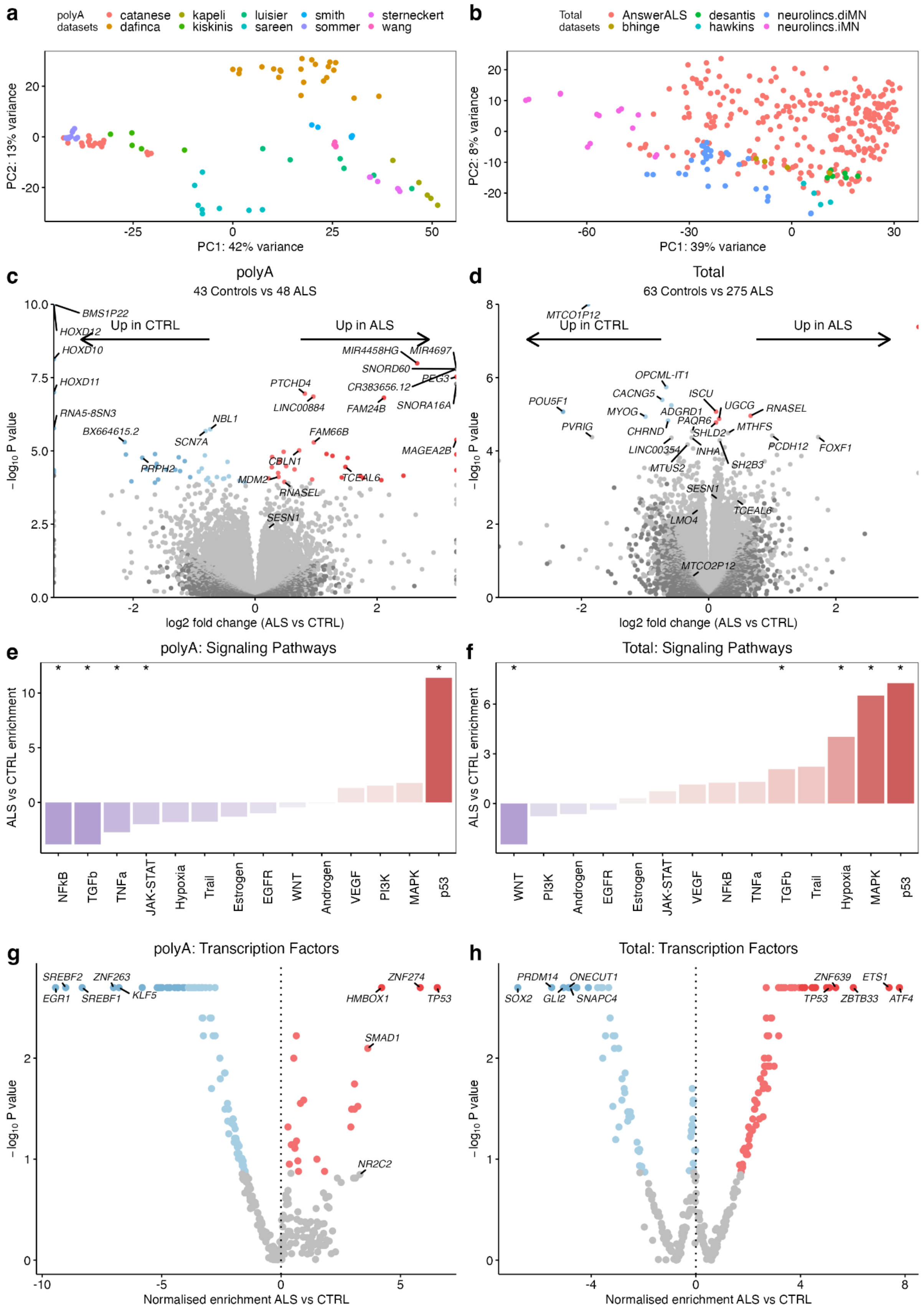
a
polyA datasets
catanese
dafinca
kapeli
kiskinis
luisier
sareen
smith
sommer
sterneckert
wang
PC1: 42% variance
PC2: 13% variance
b
Total datasets
AnswerALS
bhinge
desantis
hawkins
neurolincs.diMN
neurolincs.iMN
PC1: 39% variance
PC2: 8% variance
c
polyA
43 Controls vs 48 ALS
Up in CTRL
Up in ALS
BMS1P22
HOXD12
HOXD10
HOXD11
RNA5-8SN3
BX664615.2
SCN7A
NBL1
PRPH2
MIR4458HG
MIR4697
SNORD60
PEG3
CR383656.12
SNORA16A
PTCHD4
LINC00884
FAM24B
FAM66B
MAGEA2B
CBLN1
TCEAL6
MDM2
RNASEL
SESN1
log2 fold change (ALS vs CTRL)
$-\log_{10}$ P value
d
Total
63 Controls vs 275 ALS
Up in CTRL
Up in ALS
MTCO1P12
OPCML-IT1
CACNG5
ISCU
UGCG
RNASEL
POU5F1
MYOG
ADGRD1
PAQR6
PVRIG
CHRND
SHLD2
MTHFS
PCDH12
FOXF1
LINC00354
INHA
MTUS2
SH2B3
SESN1
TCEAL6
LMO4
MTCO2P12
log2 fold change (ALS vs CTRL)
$-\log_{10}$ P value
e
polyA: Signaling Pathways
ALS vs CTRL enrichment
NFkB
TGFb
TNFa
JAK-STAT
Hypoxia
Trail
Estrogen
EGFR
WNT
Androgen
VEGF
PI3K
MAPK
p53
f
Total: Signaling Pathways
ALS vs CTRL enrichment
WNT
PI3K
Androgen
EGFR
Estrogen
JAK-STAT
VEGF
NFkB
TNFa
TGFb
Trail
Hypoxia
MAPK
p53
g
polyA: Transcription Factors
SREBF2
ZNF263
EGR1
SREBF1
KLF5
ZNF274
HMBOX1
TP53
SMAD1
NR2C2
Normalised enrichment ALS vs CTRL
$-\log_{10}$ P value
h
Total: Transcription Factors
PRDM14
ONECUT1
SOX2
GLI2
SNAPC4
ZNF639
ETS1
TP53
ZBTB33
ATF4
Normalised enrichment ALS vs CTRL
$-\log_{10}$ P value

**Extended Data Figure 8 Sensitivity analysis of pan ALS iPSNs in poly(A) and total RNA samples separately**

a-b: Principle component analysis plots of variance stabilised transformed gene expression from polyA (a) and total (b) iPSN samples separately, coloured by dataset.

c-d: Volcano plots showing $\log_2$ fold change in differential gene expression in pan-ALS compared to control iPSNs in polyA (c) and total (d) RNA library preparation samples. Red genes are significantly (FDR < 0.05) increased and blue are decreased in ALS.

e-f: Signalling pathway activities from PROGENy showing ALS versus control iPSN normalised enrichment scores (y-axis) in polyA (e) and total (f) samples. Pathways that are increased in ALS are coloured red whilst pathways decreased are coloured blue. * represents enrichment p-value < 0.05

g-h: Transcription factor activities inferred from their respective regulon gene expression changes in ALS versus control iPSNs using DoRothEA in polyA (g) and total (h) samples. Normalised enrichment in ALS versus control (x-axis) is plotted according to the DoRothEA p-value (y-axis).

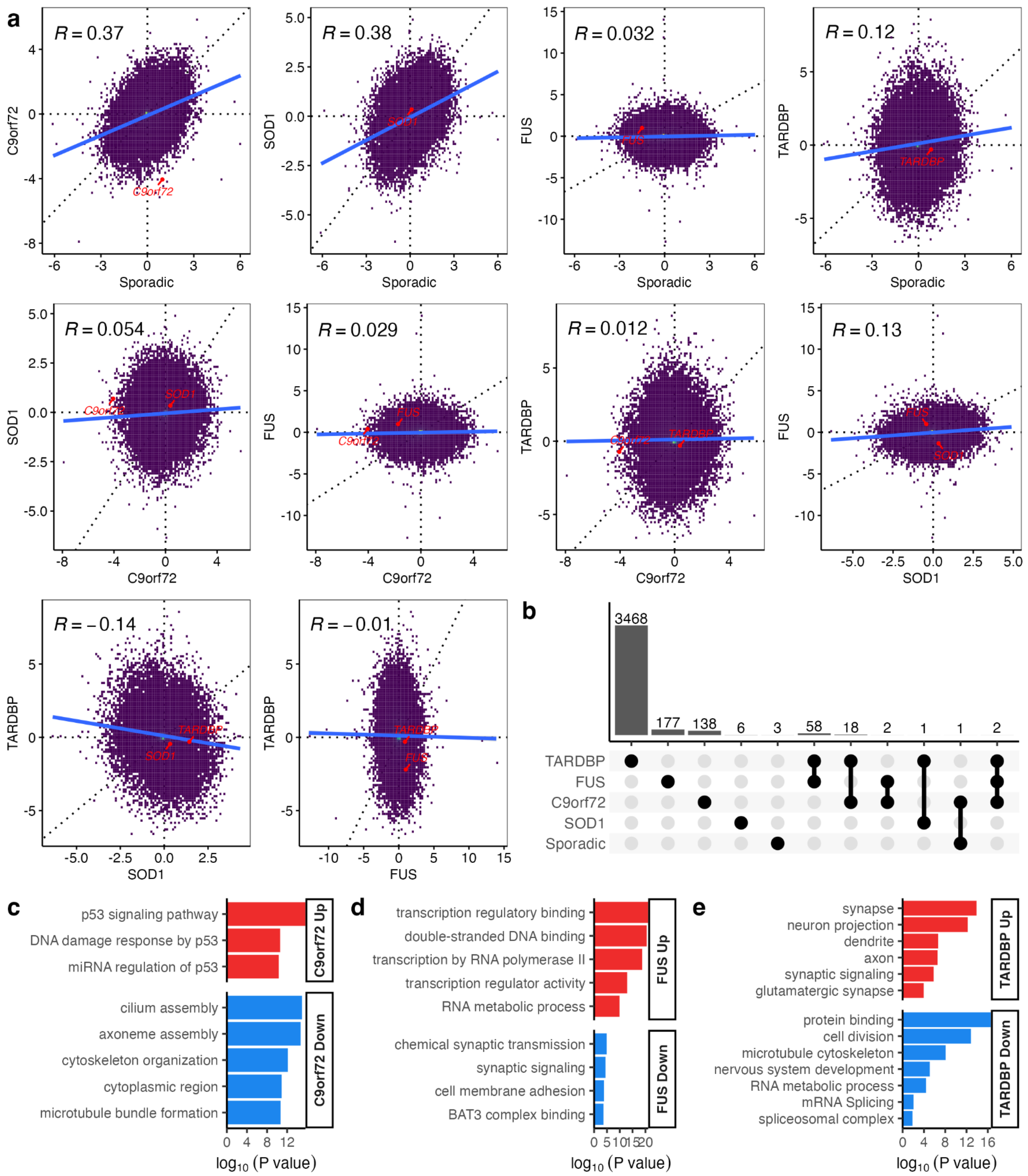


**Extended Data Figure 9 Correlating gene expression changes between genetic backgrounds**

a, Test statistics represent the sign and magnitude of the differential gene expression test for each gene compared between each mutation pair. Pearson correlation is labelled in the top left corner of each plot.

b, Upset plot showing overlapping differentially expressed genes (FDR < 0.05) between each genetic background.

c-e, Functional enrichment terms enriched in C9orf72 (c), FUS (d) and TARDBP (e).

Upregulated terms are coloured red and downregulated are blue. There were no terms enriched in SOD1 or sporadic genetic backgrounds.

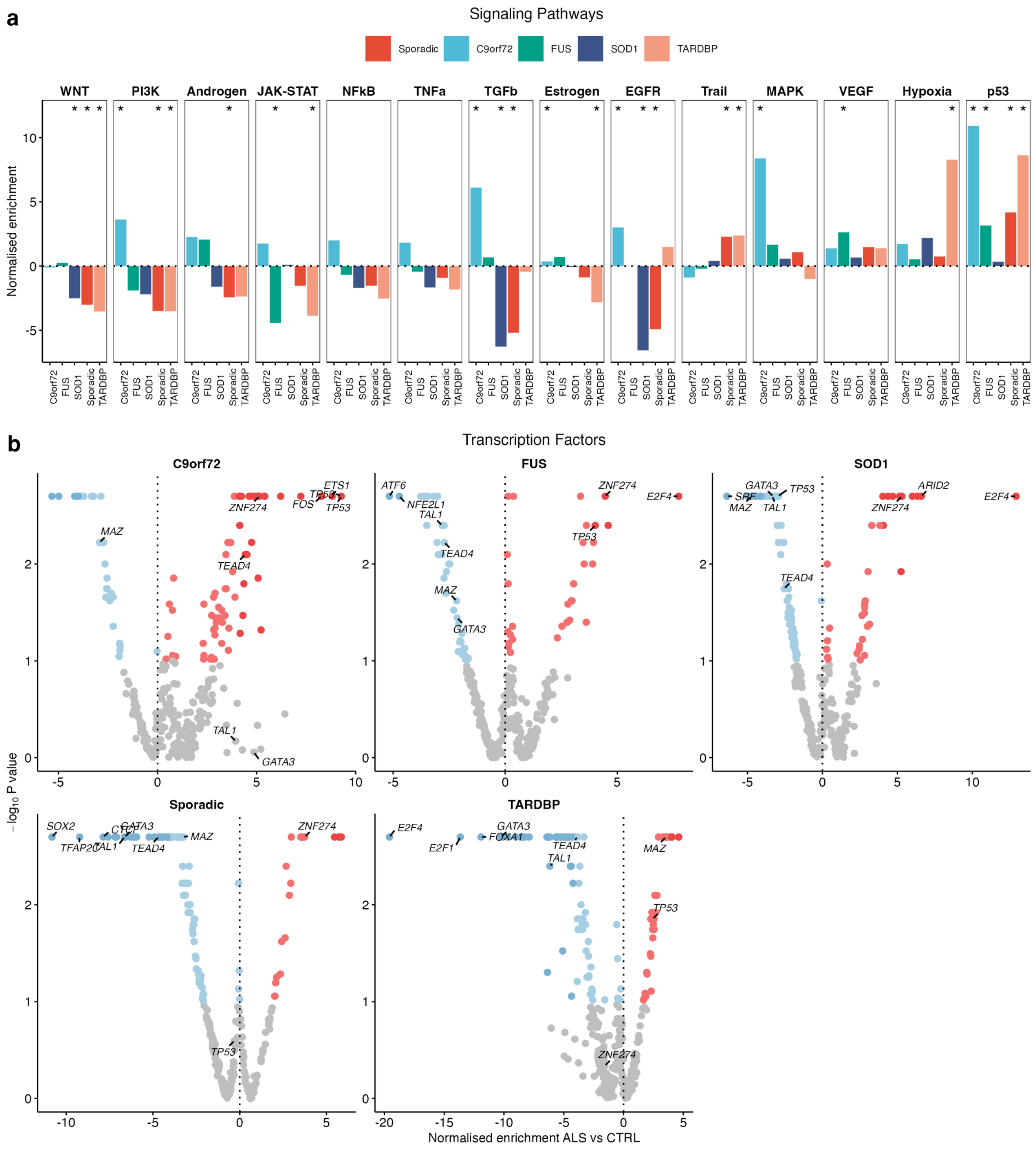


### Extended Data Figure 10 Signalling pathway and transcription factor activities between genetic backgrounds

Normalised enrichment scores in the distinct ALS genetic backgrounds versus controls for (a) PROGENy signalling pathways and (b) DoRothEA transcription factor activities. Pathways that are increased in ALS are coloured red whilst pathways decreased are coloured blue. * represent enrichment p-value < 0.05

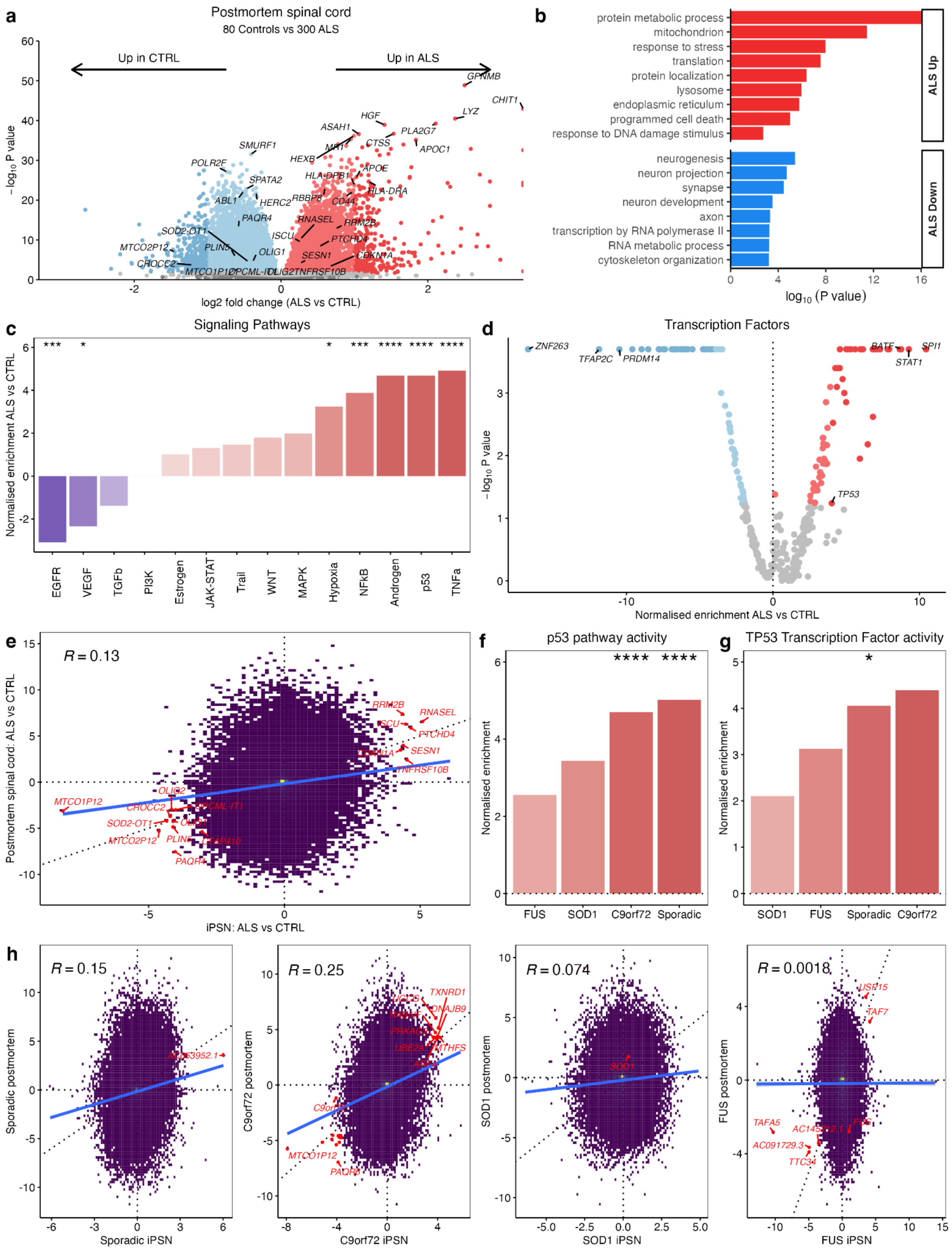


**Extended Data Figure 11 Postmortem spinal cord shows p53 activation in ALS**

a: Volcano plot of differential gene expression in ALS versus control postmortem spinal cord. Red genes are significantly (FDR < 0.05) increased and blue are significantly decreased in ALS. b, Functional enrichment terms enriched in up-regulated (red) and down-regulated (blue) differentially expressed genes in ALS versus control postmortem spinal cord. c, PROGENy signalling pathway activities in ALS versus control postmortem spinal cord. * represents enrichment p-value < 0.05. d, Transcription factor activities inferred from their respective regulon gene expression changes in ALS versus control postmortem spinal cord using DoRothEA. Normalised enrichment in ALS versus control (x-axis) is plotted according to the enrichment p-value (y-axis). e, Scatterplot of gene expression changes in ALS versus control for iPSNs (x-axis) against postmortem spinal cord (y-axis). Overlapping differentially expressed genes in both iPSNs and postmortem are coloured red. The solid blue line represents the linear correlation and Pearson correlation R = +0.083. f, p53 signalling pathway activity amongst each of the genetic backgrounds independently in postmortem spinal cord. **** represents P < 0.0001, *** P < 0.001, ** P < 0.01, * P < 0.05. g, TP53 transcription factor regulon activity in each genetic background in the postmortem spinal cord. h, Correlation of ALS versus control postmortem spinal cord (y-axis) against ALS versus control iPSN (x-axis) gene expression changes in sALS, C9orf72, SOD1 and FUS. The Wald test z-score was used as a measure of change for each gene. Overlapping differentially expressed genes are labelled as well as the mutant subgroup gene.

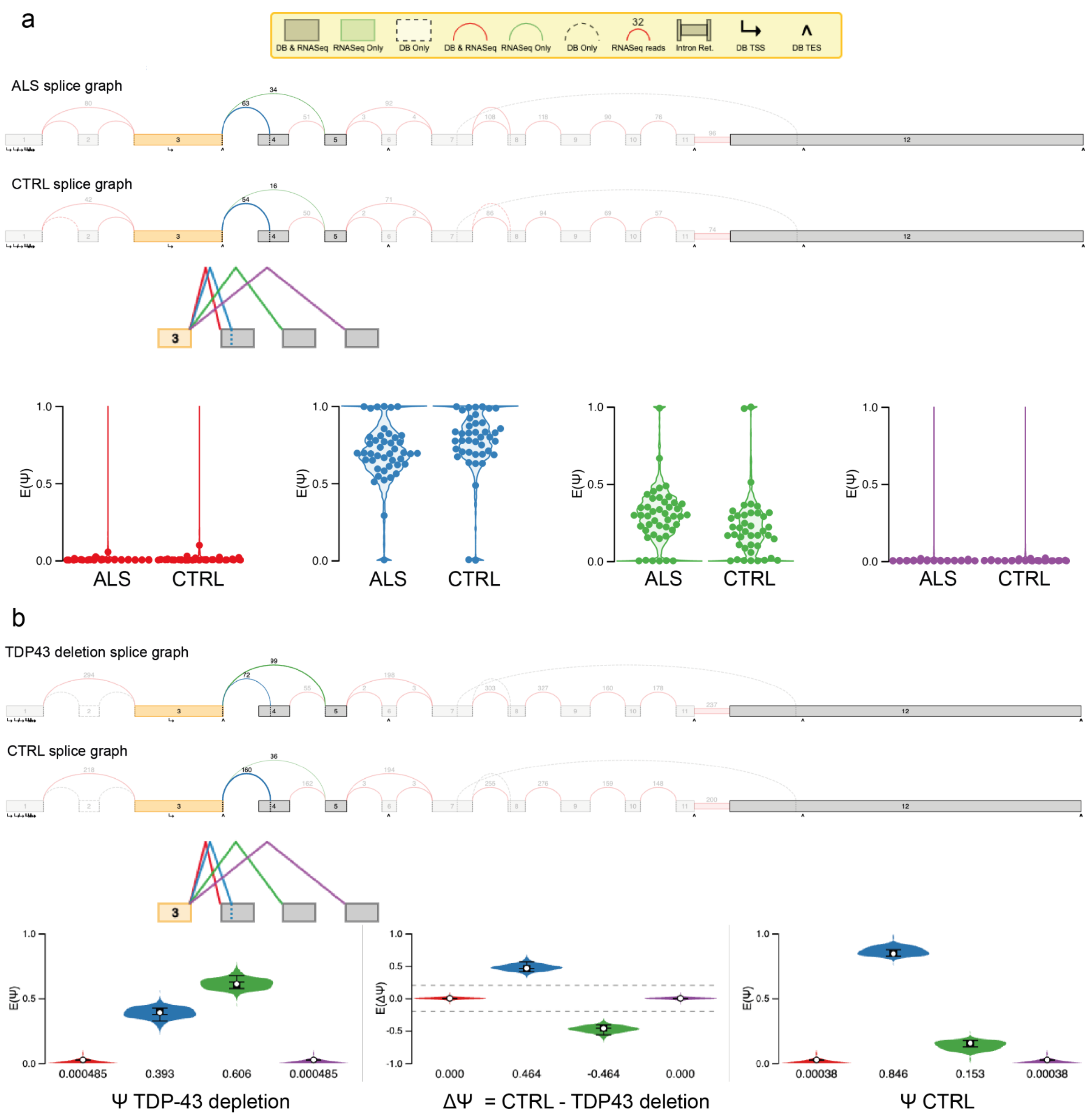


**Extended Data Figure 12 POLDIP3 splicing event in ALS iPSNs and TDP43 deletion iNeurons**

MAJIQ voila view of POLDIP3 multi-exon skipping event in (a) ALS versus control iPSNs (using heterogen function) and (b) TDP-43 deletion versus control iNeurons from Leigh-Brown et al 2022 (using deltaPSI function). In both ALS iPSNs and TDP43 deletion iNeurons there is significant inclusion of exon skipping (green variant) and exclusion of exon 4 (blue variant) at the same loci of POLDIP3.

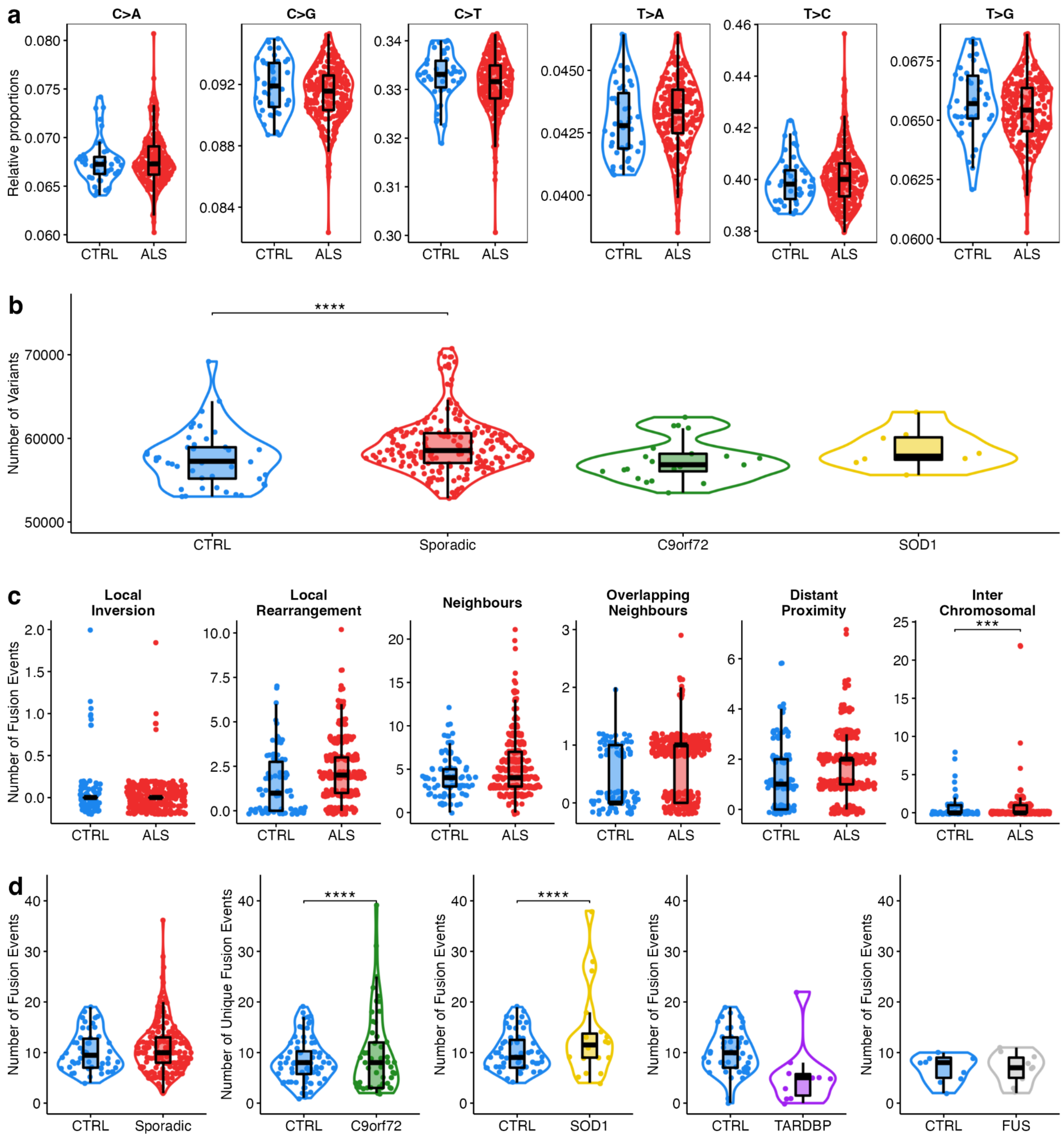


**Extended Data Figure 13 Number of RNA variants and fusions per iPSN in each genetic subgroup**

**a**, Violin plots showing the relative proportion of each base substitution type in ALS and control iPSNs. **b**, Violin plot showing the numbers of variants identified per iPSN in ALS genetic subgroups and controls samples. There were no FUS mutants and only 1 TARDBP mutant in Answer ALS. **c**, Numbers of each type of gene fusion events per iPSN in ALS (red) and CTRL (blue) samples. A generalised linear model with poisson distribution was fit, adjusting for coverage and dataset, to compare ALS with control iPSNs. **d**, Violin plots showing the numbers of gene fusion events per iPSN in each ALS genetic subgroup and controls from their respective datasets. Statistics are from the generalised linear model Wald test accounting for dataset batches and read coverage. **** $p < 0.0001$, *** $p < 0.001$, ** $p < 0.01$, * $p < 0.05$.
